# Multiple imputation of missing data under missing at random: including a collider as an auxiliary variable in the imputation model can induce bias

**DOI:** 10.1101/2023.06.16.23291497

**Authors:** Elinor Curnow, Kate Tilling, Jon E Heron, Rosie P Cornish, James R Carpenter

## Abstract

Epidemiological studies often have missing data, which are commonly handled by multiple imputation (MI). In MI, in addition to those required for the substantive analysis, imputation models often include other variables (“auxiliary variables”). Auxiliary variables that predict the partially observed variables can reduce the standard error (SE) of the MI estimator and, if they also predict the probability that data are missing, reduce bias due to data being missing not at random. However, guidance for choosing auxiliary variables is lacking. We examine the consequences of a poorly-chosen auxiliary variable: if it shares a common cause with the partially observed variable *and* the probability that it is missing (*i*.*e*. it is a “collider”), its inclusion can induce bias in the MI estimator and may increase SE. We quantify, both algebraically and by simulation, the magnitude of bias and SE when either the exposure or outcome are incomplete. When the substantive analysis outcome is partially observed, the bias can be substantial, relative to the magnitude of the exposure coefficient. In settings in which complete records analysis is valid, the bias is smaller when the exposure is partially observed. However, bias can be larger if the outcome also causes missingness in the exposure. When using MI, it is important to examine, through a combination of data exploration and considering plausible casual diagrams and missingness mechanisms, whether potential auxiliary variables are colliders.

**Contribution to the field statement:** In multiple imputation (MI), in addition to those required for the substantive analysis, imputation models often include other variables (“auxiliary variables”). Auxiliary variables that predict the partially observed variables can reduce the standard error (SE) of the MI estimator and, if they also predict the probability that data are missing, reduce bias due to data being missing not at random. We examine the consequences of a poorly-chosen auxiliary variable: if it shares a common cause with the partially observed variable *and* the probability that it is missing (*i*.*e*. it is a “collider”), its inclusion can induce bias in the MI estimator and may increase SE. We demonstrate that when the substantive analysis outcome is partially observed, the bias can be substantial, relative to the magnitude of the exposure coefficient. In settings in which complete records analysis is valid, the bias is smaller when the exposure is partially observed. However, bias can be larger if the outcome also causes missingness in the exposure. We recommmend a combination of data exploration and consideration of plausible casual diagrams and missingness mechanisms to examine whether potential auxiliary variables are colliders.

## 1. Introduction

Missing data are ubiquitous in health and social research, with multiple imputation (MI) the most flexible, general, and commonly used method for analysing partially observed datasets (1). When imputation models are appropriately specified, MI gives valid inferences if data are missing completely at random (MCAR) or missing at random (MAR), conditional on the observed data, but not (unless additional information is available) if data are missing not at random (MNAR) (Table 1). In MI, in addition to the variables used in the analysis model, imputation models often include auxiliary variables (Table 1). Auxiliary variables have two main functions: (i) to improve the predictive ability of the imputation model, over and above the information recovered via the analysis model variables, thus increasing precision (2), and (ii) to reduce bias due to data being MNAR (this is sometimes described as “making the MAR assumption more plausible”) (3). However, previous studies have shown that inclusion of auxiliary variables that are only weakly correlated with the partially observed variable, conditional on the remaining imputation model variables, can increase the standard error (SE) of the MI estimate (2, 4). In this paper, we highlight another, little known, consequence of incorrect choice of auxiliary variable: inclusion of an auxiliary variable that shares a common cause with the partially observed variable *and* its missingness (in causal inference, such a variable is referred to as a “collider” (5)) can lead to biased MI estimates by inducing a MNAR mechanism. We also demonstrate that inclusion of a collider in the imputation model may also increase SE, despite the collider being (conditionally) predictive of the missing data. The consequences of including a collider in the imputation model was discussed in principle by Thoemmes and Rose (6). Here, we quantify the bias and SE of the MI estimator based on a collider. We expand the scenarios discussed by Thoemmes and Rose, considering settings in which the (continuous or binary) partially observed variable is either the analysis model outcome or the exposure. We further illustrate our results using simulation and real data examples. All analyses were conducted using Stata (17.0, StataCorp LLC, College Station, TX). Stata code to perform the simulation studies is included in Supplementary Material, Section S8. Stata code to perform the real data analysis is included in Supplementary Material, Section S9.

**Table 1.** Missing data definitions

| **Term** | **Definition** |
| --- | --- |
| Complete Records Analysis (CRA) | Analysis is restricted to subjects who have complete data for all variables in the analysis model. |
| Missing Completely At Random (MCAR) | The probability that data are missing is independent of the observed and missing values of variables in the analysis model, and of any related variables. Data can be MCAR if missingness is caused by a variable independent of all these e.g. if missingness is for administrative reasons. |
| Missing At Random (MAR) | Given the observed data, the probability that data are missing is independent of the true values of the incomplete variable. Any systematic differences between the observed and missing values can be explained by associations with the observed data. |
| Missing Not At Random (MNAR) | If data are not MCAR nor MAR, data are said to be MNAR. The probability that data are missing depends on the (unobserved) values of the incomplete variable, even after conditioning on the observed data. |
| Multiple Imputation (MI) | MI is a method for handling missing data. It consists of three steps:<br>1. An imputation model is fitted to the observed data (this is usually some form of regression model). The missing values are replaced with draws ("imputed") from their conditional predictive distribution (after first perturbing the model parameters). This imputation stage is carried out multiple (M) times, to give M completed datasets.<br>2. The analysis model is fitted to each of the M completed datasets.<br>3. The M sets of results are combined using Rubin's rules, (23) to correctly account for the uncertainty about the missing values. |
| Auxiliary variable | A variable that is not in the analysis model but that is included as a predictor in the imputation model to recover information about the missing data. |

## 2. Bias and SE of the MI estimator including a collider in the imputation model when a continuous outcome is partially observed

### 2.1. Methods

We first consider the setting as shown in the causal diagram (or directed acyclic graph, DAG) in Figure 1. This simplified setting is chosen to give insights into the more complex settings that typically occur in epidemiological practice. We are interested in the relationship between continuous outcome (*Y*) and continuous exposure (*X*), with *β*_*YX*_ denoting the parameter of interest. Here, we assume that *X* is fully observed and *Y* is partially observed, with variable *R*_*ind*_ denoting the missingness indicator for *Y* (*R*_*ind*_ = 1 if *Y* is observed, and 0 otherwise). We assume that we know (having considered the DAG) the ‘substantive model’ we would fit to address our scientific question if there were no missing data. In this case, this is simply the regression of *Y* on *X*, because the other variables depicted in the DAG, {*Z, W, U* }, do not confound the *X*-*Y* relationship. We assume that *Z* and *W* are fully observed, and *U* denotes unmeasured variable(s).

**Figure 1.**
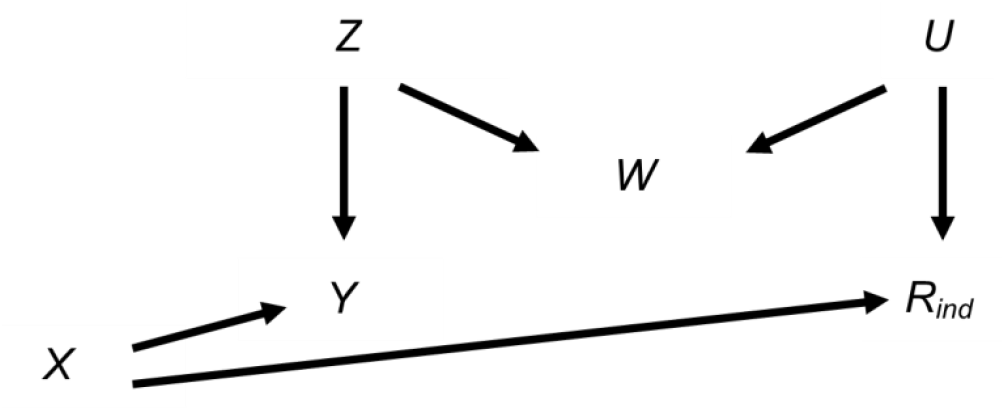
Directed acyclic graph depicting the relationship between outcome Y, exposure X, missingness indicator R_ind_, and potential auxiliary variables Z, W, and U. Lines indicate related variables, with arrows indicating the direction of the relationship; absent lines represent variables with no direct causal relation.

Since *R*_*ind*_ is unrelated to *Y* conditional on *X*, both CRA, and MI using *X* as the predictor in the imputation model for *Y*, are valid analysis strategies (7) and will yield unbiased estimates given correctly specified models. However, MI using just *X* will recover no additional information compared to CRA (8). Therefore, we may wish to include auxiliary variables in our imputation model (*i*.*e*. either *Z*, or *W*, or both) to improve the precision of our estimate of *β*_*YX*_.

For example, consider a longitudinal cohort study where we are interested in the relationship between child’s body mass index (BMI) at age 7 years (our outcome *Y*) and maternal education (our exposure *X*). In this study, suppose that BMI at age 7 years is partially observed, maternal education is fully observed, and that there are only two fully observed candidate auxiliary variables available for use in the imputation model for BMI at age 7 years: pregnancy size (singleton vs. twin birth), *Z*, and child’s birth weight, *W*, (although we note that in reality there will be many other measured and unmeasured variables related to those discussed here *e*.*g*. markers of socio-economic position). We want to choose the most appropriate set of predictors to include in the imputation model for BMI at age 7 years, choosing between: (i) maternal education, (ii) maternal education and pregnancy size, (iii) maternal education and birth weight, or (iv) maternal education, pregnancy size, and birth weight.

We assume pregnancy size is a cause of birth weight and BMI at age 7 years, but is unrelated to the missingness of BMI at age 7 years (*i*.*e*. pregnancy size is related to the other variables as depicted for *Z* in Figure 1). If we further assume that birth weight is related to missingness via some unmeasured variable(s) but is not a cause of BMI at age 7 years (*i*.*e*. birth weight is related to the other variables as depicted for *W* in Figure 1), then birth weight shares a common cause with both BMI at age 7 years and its missingness. In other words, birth weight is a collider of BMI at age 7 years and its missingness. In this case, including birth weight but not pregnancy size in the imputation model for BMI at age 7 years will induce bias in the MI estimate (in causal inference, this type of bias is often referred to as “M-bias” (9), due to the “M” shape of the causal pathways, as shown in Figure 1).

For the setting depicted in Figure 1, we provide general formulas for quantifying the bias and SE of the MI estimator when using *X* and *W*, but not *Z*, as predictors in the imputation model for *Y*. A full proof is included in the Supplementary Material (Section S2). The main arguments and results are described below.

### 2.2. Results

#### 2.2.1. Bias in the MI estimator when including a collider in the imputation model

We assume that *Y, X, Z, U*, and *W* are normally distributed, and *R*_*ind*_ is defined as follows: there exists a normally distributed variable *R* with mean *µ*_*R*_ and variance *V*_*R*_ such that 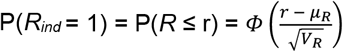, where *ϕ* denotes the cumulative distribution function of the standard normal distribution. Furthermore, we assume that each of *Y, W*, and *R* is a linear combination of the variables causing it plus an error term (with *X, Z*, and *U* having no direct causes), with no interactions, all errors uncorrelated, no model mis-specification, and no measurement error. Finally, we assume an ordinary least squares (OLS) estimator is used to obtain estimates in both analysis and imputation models.

We consider the situation in which MI is performed by replacing missing values of *Y* with draws from a linear regression model (note this is the default method for continuous variables when using *mi impute* in Stata (10) or *proc mi* in SAS (11), although predictive mean matching (12) is the default method when using *mice* in R (13)). As described above, we assume both *X* and *W* are included as predictors in the imputation model for *Y, i*.*e*. the imputation model is of the form: E(*Y*) = *α*_0_ + *α*_1_*X* + *α*_2_*W*, where E(.) denotes the expected value. Following the argument of Carpenter and Kenward (4) and noting, implicit from Figure 1, that *β*_*YX*_ conditional on *W* (*β*_*YX*|*W*_) is equivalent to *β*_*YX*_ in our scenario, the MI estimator of *β*_*YX*_ (denoted by 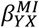) equals the regression parameter for *X* from the imputation model for *Y* based on records with observed values of *Y* (we denote this parameter by 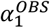). Hence, the MI estimator is unbiased only if 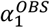 is unbiased.

In general (see Supplementary Material Section S1 for further explanation of this result), the bias of the MI estimator is bounded as follows: 0 ≤ bias ≤ |*β*_*YX*|*W,R*_ − *β*_*YX*_|. If there are no missing values of *Y*, the MI estimator is unbiased. As the probability that *Y* is missing (*i*.*e*. P(*R*_*ind*_ = 0), denoted by *π*_0_) increases, the magnitude of bias of the MI estimator increases. In the hypothetical situation in which all values are missing, bias takes its maximum value of |*β*_*YX*|*W,R*_ − *β*_*YX*_|.

#### 2.2.2. Standard error of the MI estimator when including a collider in the imputation model

The SE of the MI estimator when including collider *W* in the imputation model, 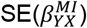, will always be greater than the SE of the imputation model coefficient 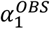, 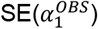, with 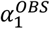 as defined above, tending towards 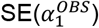 as the number of imputations increases (4). Hence, given a large number of imputations, 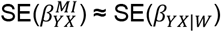 when *π*_0_ = 0 and 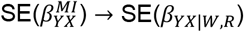 as *π*_0_ → 1 (see Supplementary Material Section S1 for further explanation of this result).

In general, the SE of the OLS estimator of a regression coefficient, SE(*β*), equals the square root of the residual variance divided by the square root of the product of the sample size (*n*) and the variance of *X* for the fitted model. Hence, we can calculate SE(*β*_*YX*|*W*_) and SE(*β*_*YX*|*W,R*_) as follows: 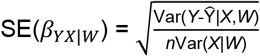 and 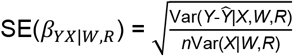, where in this setting, *n* represents the number of records with an observed value of *Y*, and Ŷ represents the mean value of *Y* predicted using the specified imputation model.

Since Cov(*X,W*) = 0 and Var(*X*|*W*) = Var(*X*) (see Supplementary Material Section S2 for proof of this and other expressions in this section), SE(*β*_*YX*|*W*_) can be expressed fairly simply as:

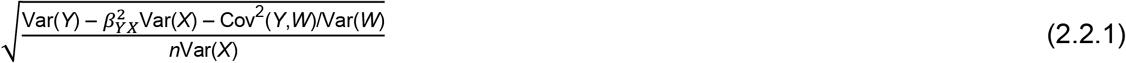

The expression for SE(*β*_*YX*|*W,R*_) is more complicated; if the imputation model parameters for *X, W*, and *R* are denoted by b_1_, b_2_, and b_3_, respectively, SE(*β*_*YX*|*W,R*_) has the general form:

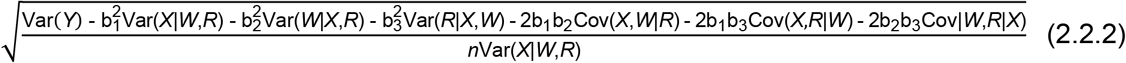

The size of this expression, relative to the magnitude of Formula 2.2.1, will depend on the strength of the associations between *Y, X, Z, W, U*, and *R*. Since Var(*X*|*W,R*) ≤ Var(*X*), if the residual variance (*i*.*e*. the numerator in Formula 2.2.2) is at least as large as that for

SE(*β*_*YX*|*W*_) (*i*.*e*. the numerator in Formula 2.2.1), SE(*β*_*YX*|*W,R*_) will be greater than SE(*β*_*YX*|*W*_) given the same sample size *n*.

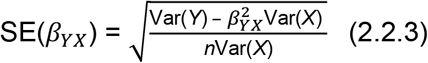

when *π*_0_ = 0,

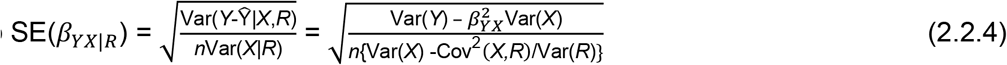

as *π*_0_ → 1 (noting *Y* is unrelated to *R* given *X* so Ŷ|*X,R* = *β*_*YX*_*X*). Note this is also, given a large number of imputations, approximately the SE of the MI estimator when only *X* is included in the imputation model. Comparing Formulas 2.2.3 and 2.2.4, we see, as expected, that SE of the CRA estimator increases as *π*_0_ → 1. Furthermore, comparing Formulas 2.2.3 and 2.2.4 with Formulas 2.2.1 and 2.2.2, the SE of the CRA estimator, or the MI estimator using only *X*, may be greater in magnitude than SE of the MI estimator including *W* in the imputation model, depending on the strength of the associations between *Y, X, Z, W, U, R*, and *π*_0_ (although the SE of the CRA estimator, or the MI estimator using only *X*, will always be greater than the SE of the MI estimator including *W* in the imputation model when *π*_0_ = 0, given Cov(*Y, W*) ≠ 0).

#### 2.2.3. Illustration of the bias and standard error of the MI estimator when including a collider in the imputation model as the proportion of missing data increases

We illustrate how the bias and SE of the MI estimator when including a collider in the imputation model vary with *π*_0_, using a simple simulation (see Supplementary Material Section S3 for further details). For reference, we also illustrate how the SE of the CRA estimator varies with *π*_0_ (the CRA estimator is always unbiased in this setting). This example is based on the relationships depicted in Figure 1, setting the mean of each variable equal to zero, all direct effect sizes equal to one, and all error variances equal to one.

Figure 2 shows, as *π*_0_ increases, (a) estimated bias and (b) estimates of SE of the MI estimator when the imputation model includes a collider, compared with SE of the CRA estimator. For reference, the true values of *β*_*YX*_, *β*_*YX*|*W,R*_, SE(*β*_*YX*|*W*_), SE(*β*_*YX*|*W,R*_), SE(*β*_*YX*_), and SE(*β*_*YX*|*R*_) are shown (with the residual variance of SE(*β*_*YX*|*W,R*_) calculated empirically due to the complexity of the algebraic form for this quantity). As expected, when there were no missing values, bias of the MI estimator equalled zero, SE of the MI estimator was equal to SE(*β*_*YX*|*W*_), and SE of the CRA estimator was equal to SE(*β*_*YX*_). As *π*_0_ increased, bias, SE of the MI estimator, and SE of the CRA estimator increased at a similar, approximately linear rate (until *π*_0_ was very close to 1), approaching |*β*_*YX*|*W,R*_ − *β*_*YX*_|, SE(*β*_*YX*|*W,R*_), and SE(*β*_*YX*|*R*_), respectively, as *π*_0_ approached 1. Bias was approximately half the maximum value when *π*_0_ = 0.5. In this particular example, for each value of *π*_0_, the SE of the MI estimator was smaller than the SE of the CRA estimator. However, note that this will not always be the case *e*.*g*. if the strength of the associations between both *Y* and *Z*, and *W* and *Z* are reduced to 0.5 (with the setting otherwise as depicted in Figure 2), SE of the MI estimator will be greater than SE of the CRA estimator if the proportion of missing data is greater than approximately 40% (see Supplementary Material Section S1, Figure S1 and also Section S5, Figure S2 which illustrates the relative precision of the MI and CRA estimators for various direct effect sizes). The difference between 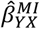 and 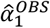 was negligible (the median difference was 0.0001, 5^th^ – 95^th^ percentile: -0.0003 – 0.0001).

**Figure 2.**
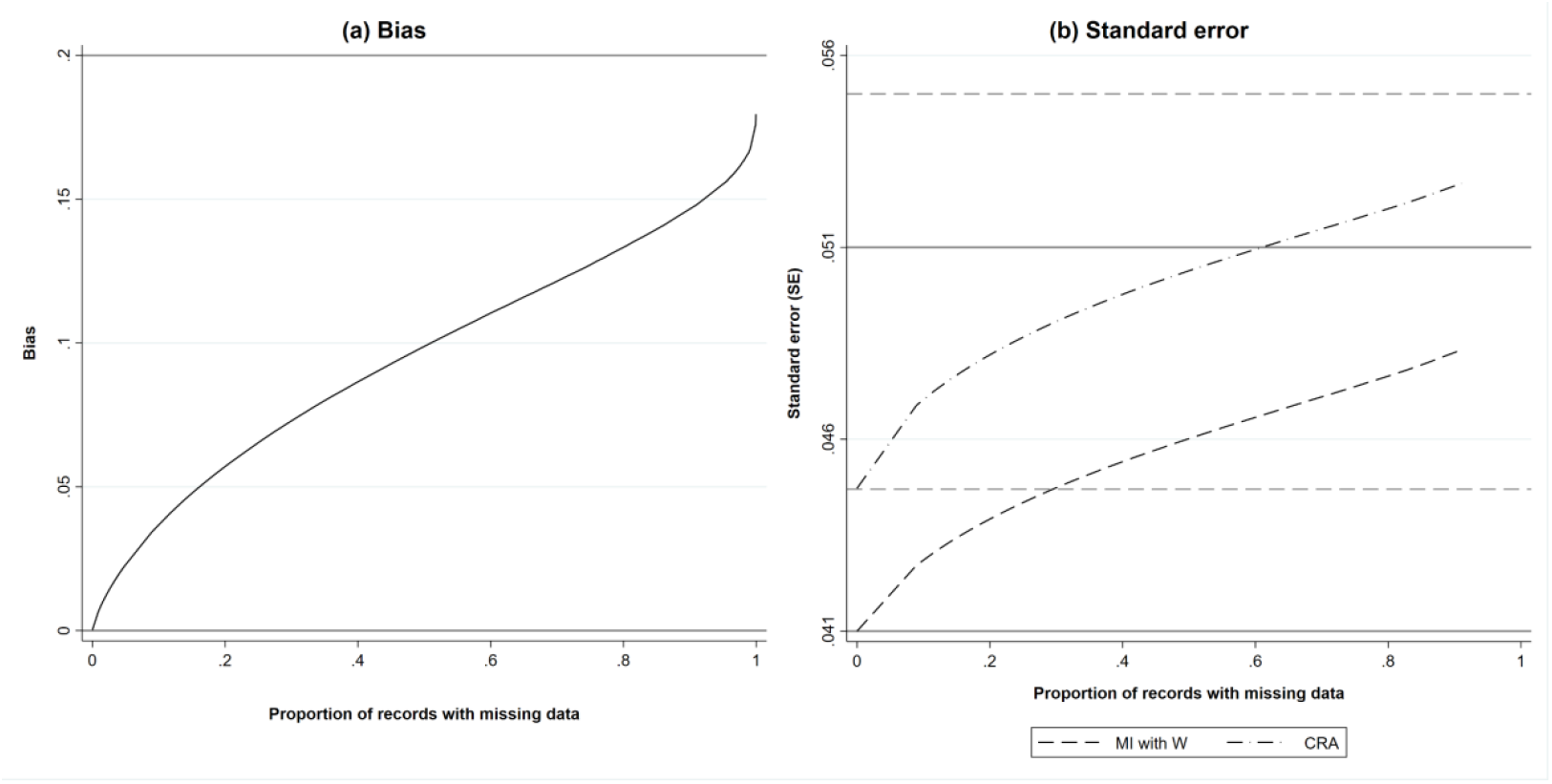
(a) Estimated bias and (b) standard error (SE) of the MI estimator of β_YX_ when the imputation model includes a collider, W, and SE of the complete records analysis (CRA) estimator of β_YX_, plotted against the proportion of records with missing data, when continuous outcome Y is partially observed, assuming 1000 observed values. All direct effect sizes and error variances equal one. Horizontal grey solid lines represent the values of bias and SE of the MI estimator when the proportion of records with missing data is zero (lower line) or tends to one (upper line). Horizontal grey dashed lines represent the values of SE of the CRA estimator when the proportion of records with missing data is zero (lower line) or tends to one (upper line).

#### 2.2.4. General expression for the maximum bias of the MI estimator when including a collider in the imputation model

In terms of the direct effect sizes and error variances, the maximum bias of the MI estimator when including a collider in the imputation model is:

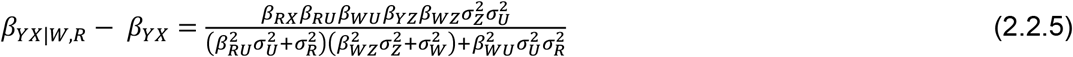

where the direct effect sizes are denoted by *β*_.._, *e*.*g. β*_*RX*_ denotes the direct effect of *X* on *R*, and the variances of the errors are denoted by *σ*^2^, *e*.*g*. 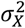 denotes the variance of the error of *X*. Formula 2.2.5 was verified by simulation (see Supplementary Material Section S4).

From Formula 2.2.5 we can see that the magnitude of the maximum bias does not depend on *β*_*YX*_ and that the direction of the maximum bias depends on the sign of the product *β*_*RX*_*β*_*RU*_*β*_*WU*_*β*_*YZ*_*β*_*WZ*_ (because 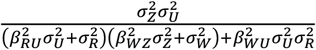 is strictly positive assuming non-zero error variances). There will be no bias if at least one of *β*_*RX*_, *β*_*RU*_, *β*_*WU*_, *β*_*YZ*_, or *β*_*WZ*_ is equal to zero, consistent with the underlying DAG (Figure 1).

#### 2.2.5. Illustration of maximum bias formula

We illustrate how the maximum bias varies with the direct effect sizes using a numerical example. In this example, we used moderate values of the direct effect sizes *β*_*RX*_, *β*_*RU*_, *β*_*WU*_, *β*_*YZ*_, and *β*_*WZ*_ (relative to the error variances 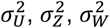, and 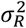, which were all equal to one): direct effect sizes were each set to 0.00, 0.25, 0.50, 0.75, or 1.00. For *β*_*RX*_ and *β*_*RU*_, note that these values correspond approximately to odds ratios (from a logistic regression model for *R*_*ind*_) of 1.00, 1.50, 2.30, 3.50, or 5.30.

Figure 3 illustrates the impact of the direct effect sizes on the maximum bias of the MI estimator. We focus particularly on the impact of *β*_*RX*_, *β*_*YZ*_, and *β*_*WZ*_ because unbiased estimates of these effect sizes can be calculated using the observed data, assuming that *X, W*, and *Z* are fully observed and - implicit from Figure 1 - that *β*_*YZ*|*R*_ = *β*_*YZ*_ (note *β*_*RU*_ and *β*_*WU*_ cannot be estimated in our setting because we assume *U* is unmeasured). In each panel, maximum bias is plotted against *β*_*YZ*_ and *β*_*WZ*_, for a single value of *β*_*RX*_ (which increases across the panels). The distribution of the maximum bias for each value of *β*_*RX*_, *β*_*YZ*_, and *β*_*WZ*_ (represented as a box-plot) is due to the variation in the other two parameters; that is, each is averaged over the values of *β*_*RU*_ and *β*_*WU*_.

**Figure 3.**
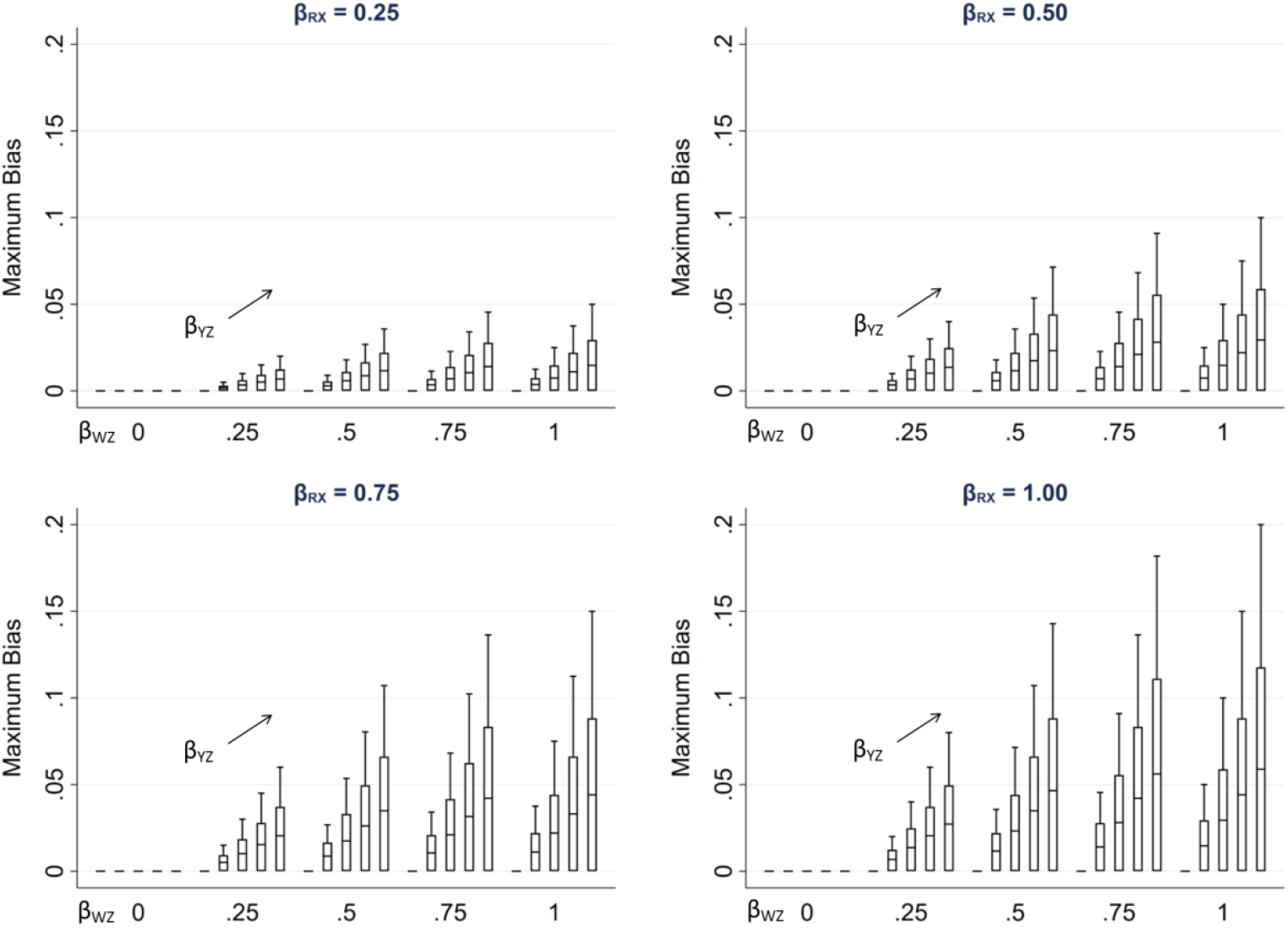
Maximum bias of the MI estimator of β_YX_ when continuous outcome Y is partially observed, varying direct effect sizes β_RX_, β_RU_, β_WU_, β_YZ_, and β_WZ_. The distribution of maximum bias in each box-plot is averaged over the values of β_RU_ and β_WU_.

As noted previously, maximum bias is equal to zero if any of the direct effect sizes are equal to zero (hence the panel with *β*_*RX*_ = 0 is not displayed), and increases with each of the direct effect parameters. Note that all parameters have a zero or positive value in this illustration. However, if, for example, we take the same parameter values as mentioned above for *β*_*RU*_, *β*_*WU*_, *β*_*YZ*_, and *β*_*WZ*_, but set *β*_*RX*_ to negative values, then the bias would be of the same magnitude but negative.

#### 2.2.6. Relative increase in precision of the MI estimator when including a collider in the imputation model

In the setting shown in Figure 1 in which bias was maximised (*i*.*e*. as *π*_0_ → 1), we also examined how the relative increase in precision of the MI estimator including *W* in the imputation model, compared with the CRA estimator, varied with the direct effect sizes. All direct effect sizes were set to 0.00, 0.50, or 1.00, and each variable had a mean of zero and an error variance of one. For each combination of direct effect sizes, SE of the CRA estimator was calculated algebraically using Formula 2.2.4. As above, due to the complexity of the expression for the SE of the MI estimator (Formula 2.2.2), this was calculated empirically. The relative increase in precision was calculated as 100 × (1 - (SE of the MI estimator)^2^/(SE of the CRA estimator)^2^). Results are illustrated in Supplementary Material Section S5, Figure S2. As discussed above, these show that, as *π*_0_ → 1, SE of the MI estimator including *W* in the imputation model can be larger or smaller than SE of the CRA estimator, depending on the magnitude of the direct effect sizes.

#### 2.2.7. Bias in other settings

We also considered the effect of collider bias in other settings. Firstly, we considered the setting in which a continuous exposure *X* was partially observed and CRA and MI were, in principle, valid, with variables related as per Figure 4. In this setting (given the same assumptions and using the same MI method as in the previous setting), the theoretical magnitude of the maximum bias (when including collider *W* in the imputation model for *X*) has a more complicated form because the imputation and substantive models are not the same. Here, the imputation model is of the form: E(*X*) = *α*_0_ + *α*_1_*Y* + *α*_2_*W*, where E(.) denotes the expected value. The MI estimator of *β*_*YX*_ will be unbiased only if an unbiased estimate of each imputation model parameter can be obtained using records with observed values of *X i*.*e*. only if 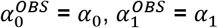, and 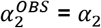.

**Figure 4.**
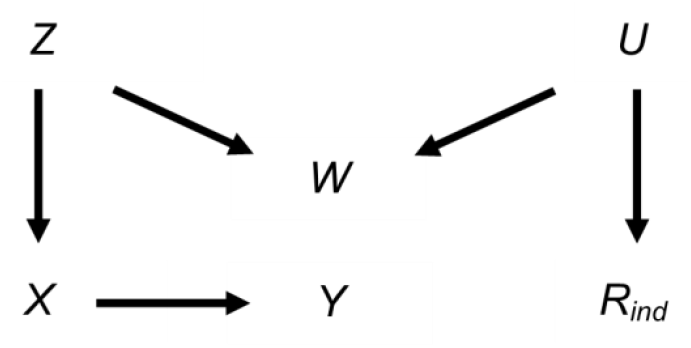
Directed acyclic graph depicting the relationship between outcome Y, exposure X, missingness indicator R_ind_, and potential auxiliary variables Z, W, and U. Lines indicate related variables, with arrows indicating the direction of the relationship; absent lines represent variables with no direct causal relation.

Taking *α*_1_ as an example, and using a similar argument to the previous setting, the bias of 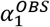 is bounded as follows: 0 ≤ bias ≤ |*β*_*XY*|*W,R*_ − *β*_*XY*|*W*_|. If there are no missing values of *X*, 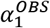 is unbiased. Bias will increase in magnitude with the probability that *X* is missing. In the hypothetical situation in which all values are missing, bias will take its maximum value of |*β*_*XY*|*W,R*_ − *β*_*XY*|*W*_|, where this depends on the magnitude of the conditional and marginal values of both the variance of *Y* and the covariance of *X* and *Y*, as well as the strength of the relationship between *W* and missingness variable *R*. Specifically, the maximum bias of 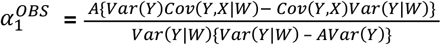, where 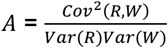 (see Supplementary Material Section S6 for further details of this derivation). Similar expressions can be derived for the maximum bias of 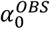 and 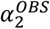.

Due to its complexity in this setting, an expression for the theoretical magnitude of the maximum bias of the MI estimator is not derived here. However, we illustrate the effect on the MI estimate from including collider *W* in the imputation model by simulation (see Supplementary Material Section S7 for further details). Note that we refer to the MI or CRA “estimate” when describing simulation study results, rather than “estimator” (which we have used when describing algebraic results). Figure 5 illustrates the impact of the direct effect sizes on the bias of the MI estimate when *X* was missing for 50% of records, focusing particularly on the impact of *β*_*YX*_, *β*_*XZ*_, and *β*_*WZ*_. In each panel, bias is plotted against *β*_*XZ*_ and *β*_*WZ*_, for a single value of *β*_*YX*_ (which increases across the panels). The distribution of the bias for each value of *β*_*YX*_, *β*_*XZ*_, and *β*_*WZ*_ (represented as a box-plot) is due to the variation in the other two parameters; that is, each is averaged over the values of *β*_*RU*_ and *β*_*WU*_. Figure 5 shows that bias is very small, regardless of the direct effect sizes. In addition, examining the relative increase in precision, compared with the CRA estimate (see Supplementary Material, Section S7, Figure S3), shows that the SE of the MI estimate including *W* in the imputation model can be larger or smaller than SE of the CRA estimate, depending on the magnitude of the direct effect sizes.

**Figure 5.**
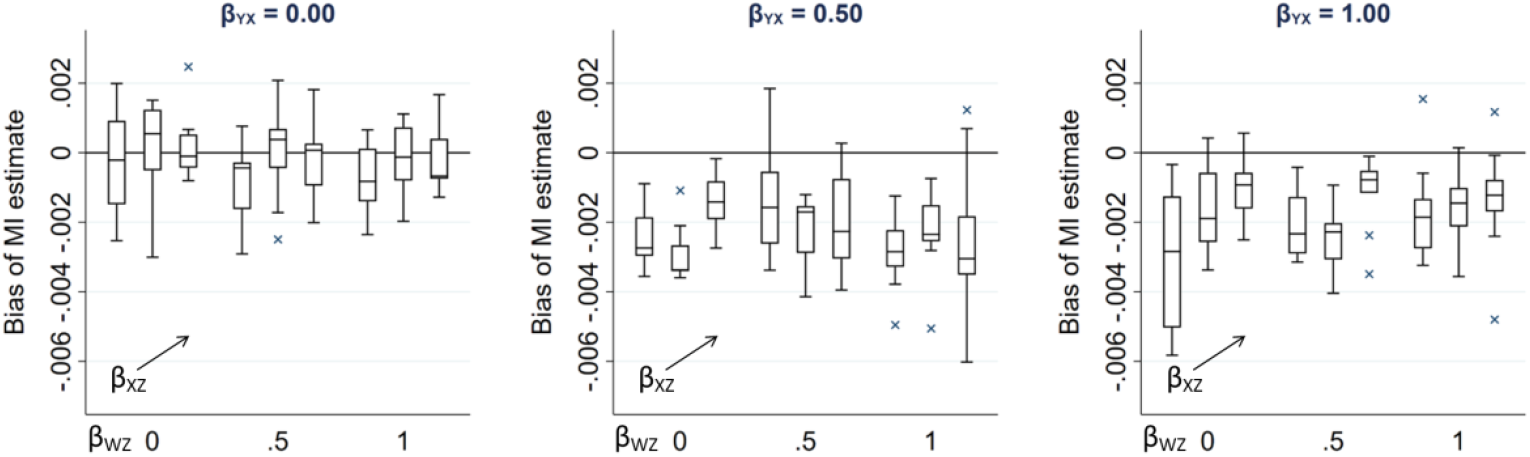
Bias of the MI estimate of β_YX_ when 50% of values of a continuous exposure X are missing, varying direct effect sizes β_YX_, β_XZ_, β_WZ_, β_RU_, and β_WU_. The distribution of bias in each box-plot is averaged over the values of β_RU_ and β_WU_.

In similar settings with a binary partially observed variable (*i*.*e*. the same settings as depicted in Figures 1 and 4 but with either partially observed binary *Y* or partially observed binary *X*), the bias of MI estimates will be approximately the same magnitude as for the continuous cases, provided the probability of each value of the binary variable is not close to 0 or 1 (see Supplementary Material Section S7, Figures S4-5). This follows in each case by assuming that the binary variable has an underlying normal distribution, in which case the results described here will still approximately apply.

#### 2.2.8. Bias when missingness of the exposure additionally depends on the outcome

In our setting with a partially observed continuous exposure *X*, the magnitude of bias was much smaller than in the setting with a partially observed continuous outcome *Y*. This is because there is only one pathway between the partially observed variable and its missingness in the *X* setting (via *Z*-*W-U*), whereas there are two pathways in the *Y* setting (via *Z*-*W-U* and *X*). Hence, the cumulative bias (*i*.*e*. the sum of the bias via each pathway) is potentially larger in the *Y* setting. Therefore, to provide a more comparable setting to that when *Y* is partially observed, we considered an additional setting when continuous variable *X* was partially observed, in which *Y* was also a cause of missingness of *X* (Figure 6). The relationships depicted in Figure 6 are the same as those in Figure 4, with the addition of an arrow from *Y* to *R*. There are now two potential pathways between *X* and its missingness, via *Z*-*W-U* and *Y*. Note that CRA is no longer valid in this setting, because missingness depends on the analysis outcome *Y*. However, MI using *Y*, or *Y* and *Z*, in the imputation model for *X* would be valid. Using the same simulation approach as before (see Supplementary Material Section S7 for further details), Figure 7 illustrates the effect on the MI estimator from including collider *W* in the imputation model. Figure 7 shows that when missingness in *X* is caused by *U* and *Y* and *β*_*YX*_ is close to 0, bias is similar in magnitude to that in the setting in which missingness in *Y* is caused by *U* and *X*.

**Figure 6.**
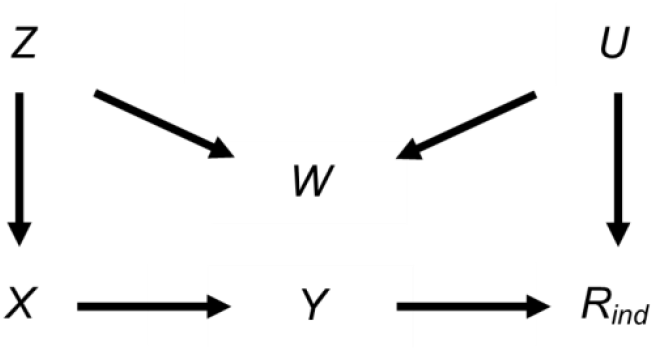
Directed acyclic graph depicting the relationship between outcome Y, exposure X, missingness indicator R_ind_, and potential auxiliary variables Z, W, and U. Lines indicate related variables, with arrows indicating the direction of the relationship; absent lines represent variables with no direct causal relation.

**Figure 7.**
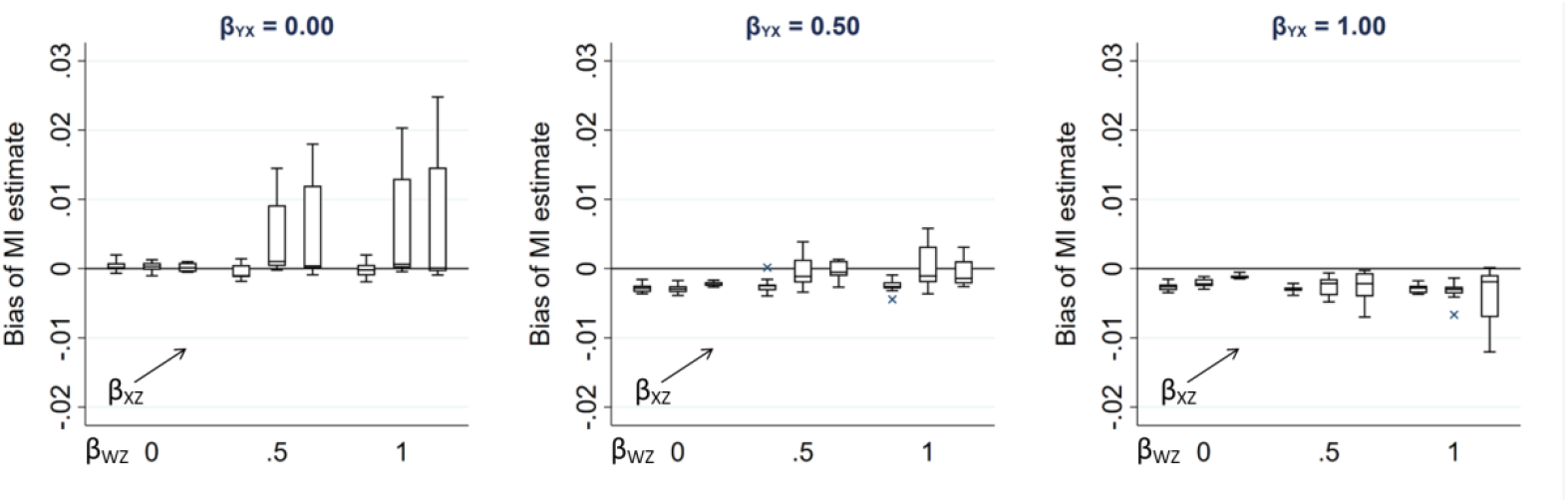
Bias of the MI estimate of β_YX_ when 50% of values of a continuous exposure X are missing, varying direct effect sizes β_YX_, β_XZ_, β_WZ_, β_RU_, and β_WU_. The distribution of bias in each box-plot is averaged over the values of β_RU_ and β_WU_.

## 3. Real data example

### 3.1. Methods

We illustrate use of our formula for maximum bias given a partially observed continuous outcome using data from the Avon Longitudinal Study of Parents and Children (ALSPAC). ALSPAC is a prospective study which recruited pregnant women with expected dates of delivery between 1^st^ April 1991 and 31^st^ December 1992, in the Bristol area of the UK (14, 15). We used data from the initial recruitment phase, in which 14,541 pregnant women enrolled, resulting in 14,062 live births (13,988 alive at one year of age). Children and their mothers have been followed up since birth through questionnaires, clinics, and linkage to routine datasets. Ethical approval for the study was obtained from the ALSPAC Ethics and Law Committee and local research ethics committees. Informed consent for the use of data collected via questionnaires and clinics was obtained from participants following the recommendations of the ALSPAC Ethics and Law Committee at the time.

Here, our substantive model of interest was the regression of child’s body mass index at age 7 years (*bmi7*) on maternal education (*mated*: a binary variable indicating whether the child’s mother held a post-16 years qualification). We restricted analysis to all singletons and first-born twins (excluding the second-born twin to avoid family-level clustering) who were alive at one year (n = 13,745). For illustrative purposes, we assumed that the exposure and auxiliary variables were fully observed (in reality, a small proportion of participants had missing values for these variables: 1684 participants, 12%, were missing values of *mated*, n = 1510, *bwt*, n = 150, or both, n = 24) and that there were only two candidate auxiliary variables available for use in the imputation model for *bmi7*: pregnancy size (*pregsize*: singleton vs. twin birth), and child’s birth weight (*bwt*) (in reality, a large amount of individual-level data are available: the ALSPAC study website contains details of all available data through a fully searchable data dictionary and variable search tool: http://www.bristol.ac.uk/alspac/researchers/our-data/). Therefore, we analysed 12,061 participants with observed values of *mated, pregsize*, and *bwt*, of whom 7248 (60%) had an observed value of *bmi7*.

Figure 8 depicts the relationships, based on prior research (16-19), between *bmi7, mated, pregsize, bwt*, and missingness indicator *R*_*ind*_ (a binary variable indicating whether *bmi7* is observed), plus unmeasured variable(s), *U* (related to the analysis model variables and/or their missingness *e*.*g*. markers of socio-economic position, SEP). Lines indicate related variables, with arrows indicating the direction of the relationship; absent lines represent variables with no direct causal relation. Straight, solid lines depict the relationships assumed in the theoretical scenario in which *Y* is MAR; curved, dashed lines depict additional relationships that are plausible in our real data example. For example, in the theoretical scenario, we assume that only *X* and *Z* cause *Y*, and only *X* and *U* cause missingness in *Y*. In the real data scenario, it is plausible that *bmi7* is MNAR, because *U* may be related to both *bmi7* and *R*_*ind*_. We assume that *pregsize* is not a cause of *R*_*ind*_, although *pregsize* may be related to *R*_*ind*_ via *U* (*e*.*g*. because assisted reproduction is associated with higher SEP). Similarly, we assume that *bwt* is not a cause of *bmi7* or *R*_*ind*_, but shares a common cause with both *bmi7* and *R*_*ind*_ *i*.*e. bwt* is a collider.

**Figure 8.**
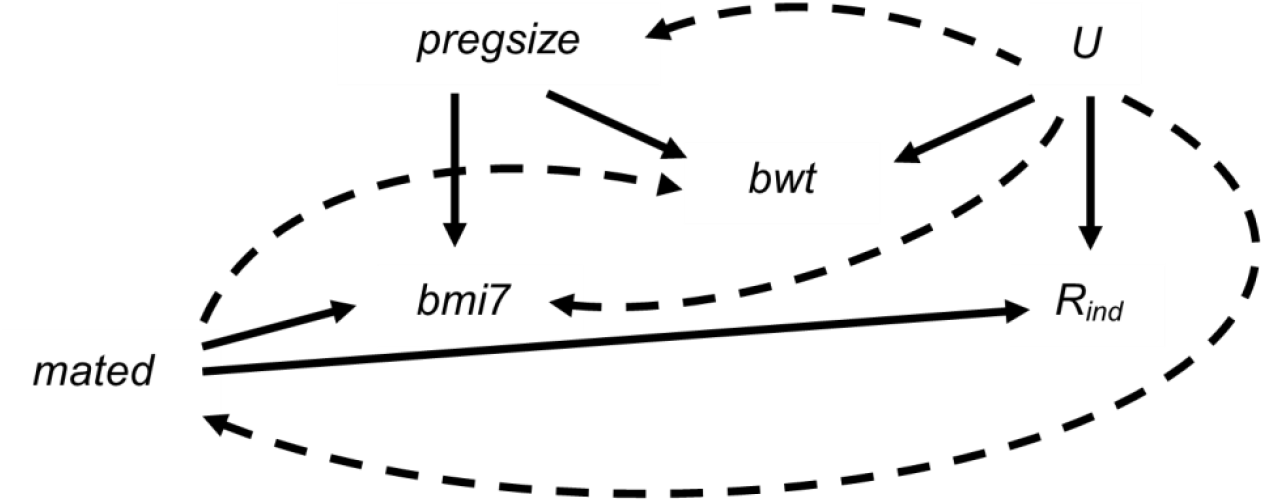
Directed acyclic graph depicting the relationship between child’s body mass index at age 7 years (bmi7), maternal education (mated: a binary variable indicating whether the child’s mother held a post-16 years qualification), pregnancy size (pregsize: singleton or twin birth), child’s birth weight (bwt), missingness indicator R_ind_ (a binary variable indicating whether bmi7 is observed), and unobserved variable(s) U. Lines indicate related variables, with arrows indicating the direction of the relationship. Straight solid lines depict the relationships assumed in the theoretical scenario in which the analysis model outcome is missing at random; curved dashed lines depict additional relationships that are plausible in our real data example; absent lines represent variables with no direct causal relation.

We assessed the potential impact on the MI estimate from including a collider (*bwt*) in the imputation model for *bmi7* in two steps:

1. We assessed whether our hypothesised relationships with *bwt* were realistic by exploring the relationships between *mated, pregsize, bwt*, and *R*_*ind*_. We assessed relationships using linear or logistic regression models (for continuous and binary outcomes, respectively), for each pair of variables in turn (deciding which variable was the dependent variable and which the explanatory variable in any given pair according to the probable causal direction), adjusting for any observed confounders.
2. Based on our results from Step 1, we applied our formula (Formula 2.2.5) for maximum bias of the MI estimator if hypothesised collider *bwt* was included in the imputation model for *bmi7*. Since not all the direct effect sizes were estimable from the observed data, we used an alternative (equivalent) version of our maximum bias formula, expressed in terms of the variances and covariances of the observed (or partially observed) variables. We also assumed (without loss of generality) that *R* had a mean of zero and a variance of one. Therefore, we used the following version of the formula to calculate maximum bias: 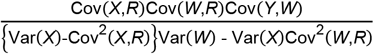

where, in our setting, *X* denotes *mated, W* denotes *bwt*, and *Y* denotes *bmi7*. Since we observe *R*_*ind*_ (*i*.*e*. whether or not *bmi7* is observed) rather than the underlying normal variable *R*, covariance terms involving *R* were approximated by applying the general rule for transforming a parameter from a logistic to a probit model (20) (valid unless the proportion of complete records is very close to 0 or 1): 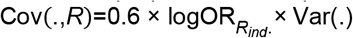, where 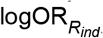. denotes the logarithm of the odds ratio (*i*.*e*. the regression parameter) from a logistic regression of *R*_*ind*_ on the specified covariate. For example, Cov(*X, R*) was approximated by 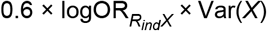. We estimated Var(*X*) using the normal approximation to the binomial because *X* was binary. We estimated Cov(*Y, W*) using the complete records and other terms using all records. For simplicity, we assumed that the relationship between *bwt* and *bmi7* was linear. We also assumed that estimates of the variances and covariances used in our maximum bias formula were unbiased (which may not have been the case if *Y* was MNAR or if there were unmeasured confounders).

We compared our estimate of the exposure coefficient based on our formula for maximum bias to both the CRA estimate and MI estimates using no auxiliary variables or either *pregsize*, or *bwt*, or both, as auxiliary variables. Each imputation model also included the analysis model exposure, *mated*. We used a large number of imputations (100) to ensure we obtained stable estimates of the exposure coefficient and its SE.

### 3.2. Results: magnitude of bias due to a collider auxiliary variable

#### Step 1

Relationships between *mated, pregsize, bwt*, and *R*_*ind*_ are summarised in Table 2. In particular, these suggest that *R*_*ind*_ is strongly associated with both *mated* and *bwt*, but less so with *pregsize*. However, adjusting for *bwt* increases the strength of the relationship between *R*_*ind*_ and *pregsize* (unadjusted odds ratio, OR, 1.07, 95% confidence interval, CI, 0.77 - 1.48 vs. adjusted OR 1.25, 95% CI 0.90 - 1.75). These results, combined with our prior knowledge of the data, suggest that *bwt* is a collider. Therefore, inclusion of *bwt* in the imputation model for *bmi7* may induce or inflate bias due to data being MNAR.

**Table 2.**
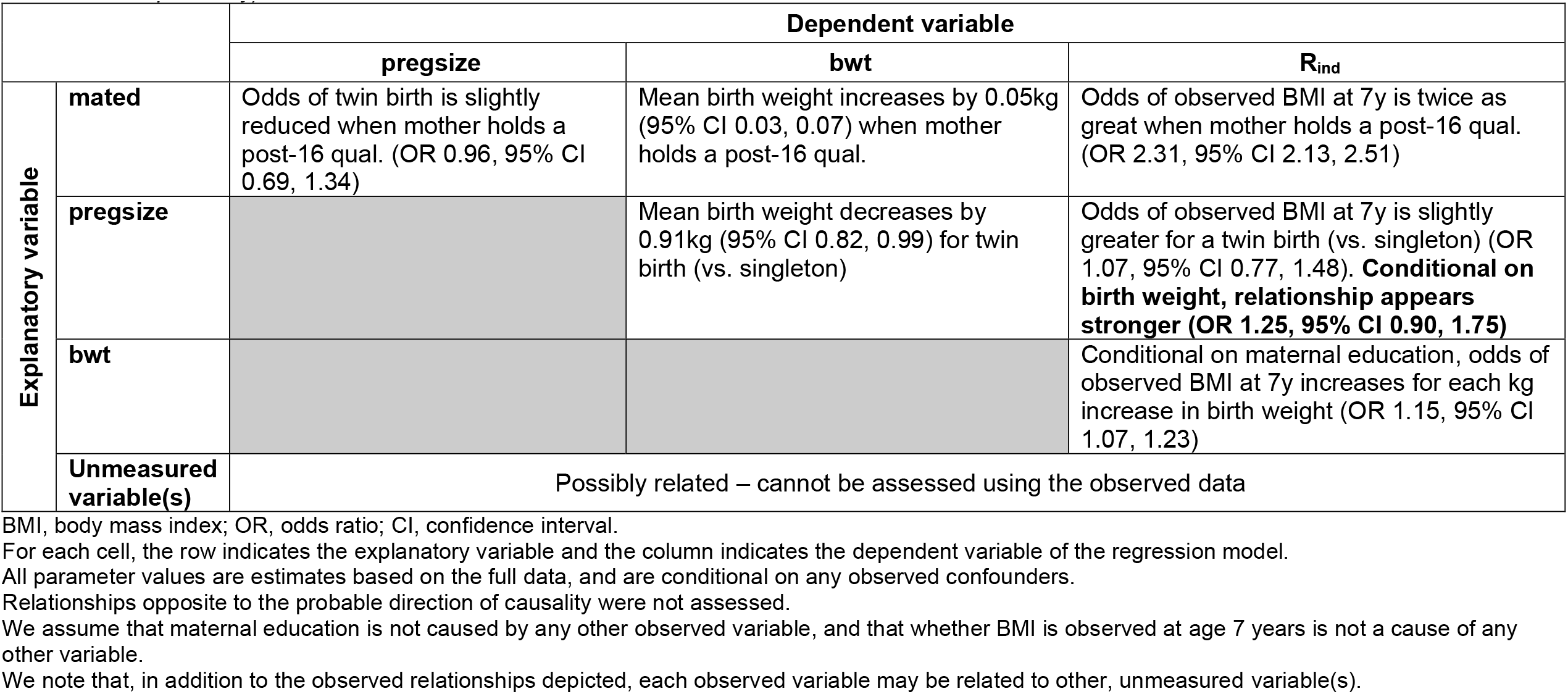
Relationships between maternal education (mated), pregnancy size (pregsize), child’s birth weight (bwt), and whether child’s body mass index (BMI) was observed at age 7 years (R_ind_), determined using linear or logistic regression models (for continuous and binary outcomes, respectively).

#### Step 2

Substituting values based on the observed data (as per Table 2, and additionally, using estimates of Var(*W*), Var(*X*), and Cov(*Y,W*) of 0.286, 0.228, and 0.171, respectively) into our theoretical expression, we estimated the maximum bias from including *bwt* in the imputation model for *bmi7* to be 0.008 (towards the null). This result suggests that even though there is the possibility of collider bias due to inclusion of *bwt* in the imputation model, the magnitude of bias is small in this particular setting.

Analysis results (Table 3) confirmed that CRA and MI estimates of the exposure coefficient were very similar, regardless of the auxiliary variable(s) used in the MI procedure. However, as predicted, there was slight attenuation in the MI estimate when *bwt* was included in the imputation model for *bmi7*. This was the case even when *pregsize* was also included. This suggests that there was at least one other unobserved variable that had similar relationships with other variables as *pregsize* (*e*.*g*. child sex), so adjusting for *pregsize* did not completely remove the bias induced by inclusion of *bwt* in the imputation model. The difference between the CRA estimate and the MI estimate including *bwt* was 0.023 (towards the null), which was larger than our estimate based on the theoretical magnitude of bias, although in the same direction.

**Table 3.** Mean change in child’s body mass index (kg/m^2^) at age 7 years when mother holds a post-16 qualification (vs. no post-16 qualification), estimated using different analysis strategies

| **Analysis strategy** | **Estimate (SE)** | **95% CI** |
| --- | --- | --- |
| Complete records analysis | -0.108 (0.049) | -0.203, -0.013 |
| MI with no auxiliary variables | -0.106 (0.049) | -0.209, -0.011 |
| MI with pregnancy size as auxiliary variable | -0.107 (0.047) | -0.198, -0.015 |
| MI with child's birth weight as auxiliary variable | -0.085 (0.047) | -0.176, 0.007 |
| MI with pregnancy size and child's birth weight as auxiliary variables | -0.091 (0.050) | -0.189, 0.006 |

SE, standard error; CI, confidence interval.

As expected, the SE of the CRA estimate was similar to the SE of the MI estimate using no auxiliary variables and larger than the SE for MI estimates using *pregsize* or *bwt* as auxiliary variables. However, the SE of the MI estimate using both *pregsize* and *bwt* as auxiliary variables was larger than for all other analysis strategies. This may be because *pregsize* has only a weak direct effect on *bmi7, i*.*e. pregsize* is largely redundant if the imputation model already includes *bwt*; thus its addition leads to a decrease in precision (4).

## 4. Discussion

In this paper, we quantify, algebraically and by simulation, the magnitude of bias and SE of the MI estimator induced by including a collider in the imputation model, in settings where it is possible to specify an imputation model that gives unbiased inference for the population parameter values. We have derived an algebraic expression for the maximum bias and its relationship to the proportion of incomplete records when a continuous outcome is partially observed. We have demonstrated that in this setting (and also if the outcome is binary), the bias can be substantial, relative to the magnitude of the exposure coefficient. We found, in settings in which CRA was valid, the bias due to inclusion of a collider in the imputation model was smaller when the exposure in the analysis model (either binary or continuous) was partially observed. However, bias was larger in magnitude if the outcome also caused missingness in the exposure (in which case CRA was no longer valid but MI, using a correctly specified imputation model and correct choice of auxiliary variables, was valid).

When the outcome is partially observed, we have shown that the magnitude of the bias of the MI estimator from including a collider in the imputation model depends on the magnitude of the associations between the exposure and missingness, between the collider and missingness, and between the collider and the outcome, as well as on the proportion of missing data. Crucially, it does not depend on the magnitude of the association between outcome and exposure. Therefore, if the association between outcome and exposure is much weaker than the associations between other pairs of variables and the proportion of incomplete records is fairly large (precisely the situation in which one may wish to use auxiliary variables), the relative bias of the MI estimator could be substantial. In addition, since the direction of bias depends on the signs of the associations between other pairs of variables (and not on the sign of the association between outcome and exposure), it is possible, for example, that this could bias the estimator to the extent that a weak positive association is incorrectly estimated as a stronger negative association.

In our real data example, we assumed that both auxiliary variables (direct predictor pregnancy size and collider birth weight) were measured. However, note that the bias can still be estimated even if the direct predictor is unmeasured, because the maximum bias formula does not depend on this variable. However, in this case, assessing whether an auxiliary variable is a collider may need to rely on both prior knowledge and inspection of the hypothetical causal model of interest, because it may be difficult to assess whether it is a collider using the observed data alone. The likely impact of including a collider in the imputation model(s) can still be assessed using our suggested formula and/or our plots based on simulations, estimating the strength of each relevant association using either the observed data or published results.

In addition to inducing bias, including a collider in the imputation model may increase, rather than decrease, the SE of the MI estimator. We have shown that this depends on the magnitude of the associations between the exposure, outcome, collider, and missingness. However, inclusion of a collider in the imputation model may recover more information about the missing data than CRA, or MI including only the other analysis model variables in the imputation model, and increase precision. Therefore, where the likely bias from inclusion of a collider is sufficiently small, we recommend performing a sensitivity analysis, comparing the precision of the MI estimate when the imputation model does or does not include a collider. If the gain in precision is sufficiently large, it may be preferable to include a collider in the imputation model, at the expense of some bias, especially if no other auxiliary variables are available.

A strength of our approach is that we have considered a range of scenarios, in which the partially observed variable is either the analysis model outcome or the exposure, as well as either continuous or binary. By using both algebraic quantification and simulation, we have been able to provide a detailed illustration of the effect on both bias and SE, and how these are related to the magnitude and sign of individual assocations between exposure, outcome, auxiliary variables, and missingness. A limitation of our study is that in each of our scenarios, only a single variable has missing values. When multiple variables have missing values, assessing whether imputation models include colliders is likely to be a more complex process. However, we would expect our findings to extend to these situations.

In summary, we conclude that, although auxiliary variables have the potential to improve precision of the MI estimate and reduce bias compared with an imputation model that only includes analysis model variables, poorly-chosen auxiliary variables can increase both bias and SE. Therefore, it is important that auxiliary variables are selected carefully. In particular, we recommend examining whether any potential auxiliary variables are colliders. This can be achieved through a combination of data exploration and consideration of the plausible casual diagrams and missingness mechanisms (*e*.*g*. by using a missingness DAG (21, 22)).

## Data Availability

Stata code to verify theoretical results, and also to generate and analyse the data as per the simulation studies is included in Supplementary Material, Section S8. Stata code to analyse the real data example is included in Supplementary Material, Section S9. The real data are not publicly available due to privacy restrictions.

## Acknowledgements

We are extremely grateful to all the families who took part in the ALSPAC study, the midwives for their help in recruiting them, and the whole ALSPAC team, which includes interviewers, computer and laboratory technicians, clerical workers, research scientists, volunteers, managers, receptionists and nurses.

