## Supplementary Material for "Multiple imputation of missing data under missing at random: including a collider as an auxiliary variable in the imputation model can induce bias"

*Section S1. Derivation of the MI estimator when a continuous outcome Y is partially observed and the imputation model includes collider variable W*

Recall from the main text that the MI estimator, $\beta_{YX}^{MI}$, equals the regression parameter for *X* from the imputation model for *Y* ($\text{E}\left( \text{Y} \right)\text{ }\text{=}\text{ }\alpha_{0}\text{ }\text{+}\text{ }\alpha_{1}\text{X}\text{ }\text{+}\text{ }\alpha_{2}\text{W}$) based on records with observed values of *Y* (we denote this parameter by $\alpha_{1}^{OBS}$). We can express $\alpha_{1}^{OBS}$ in terms of the missingness indicator, $\text{R}_{\text{ind}}$, such that $\alpha_{1}^{OBS} = \beta_{YX|W,R_{ind}=1}$, where the probability that $\text{R}_{\text{ind}}$ equals 1, P($\text{R}_{\text{ind}}$ = 1), is denoted by $\pi_{1}$ and P($\text{R}_{\text{ind}}$ = 0) is denoted by $\pi_{0}$ = ($1-\pi_{1}$). As per the main text, we assume there exists a normally distributed variable *R* with mean $\mu_{R}$ and variance $V_{R}$ such that $\pi_{1}$ = P($\text{R}_{\text{ind}}$ = 1) = P(*R* ≤ r) = $\Phi\left( \frac{r - \mu_{R}}{\sqrt{V_{R}}} \right)$, where $\Phi$ denotes the cumulative distribution function of the standard normal distribution. Here we assume, without loss of generality, that *R* has a marginal standard normal distribution *i.e.* $\mu_{R}$ = 0 and $V_{R}$ = 1.

Then $\alpha_{1}^{OBS} \text{=} \beta_{YX|W,R_{ind}=1}$ $\text{=}\text{ }\beta_{YX|W,R \leq r}$ represents the average value of $\beta_{YX|W,R}$ for all values of *R* ≤ *r* where *r* = $\Phi^{-1}\left( \pi_{1} \right)$, *e.g.* if $\pi_{1}$ = 0.5, $\alpha_{1}^{OBS}$ is the average value of $\beta_{YX|W,R}$ for all *R* ≤ 0. Since *R* is normally distributed, taking values across the range $(-\infty, \infty)$, we can write this as: $\alpha_{1}^{OBS} = \beta_{YX|W,R_{ind}=1} = \int_{-\infty}^{r} \beta_{YX|W,R=s} p(s)ds$ where $p(s)$ denotes the probability that *R* = *s* given *s* $\in(-\infty, \text{r}]$.

In general, $p(s)$ = P(*R* = *s* | *R* ≤ *r*) = P(*R* = *s* & *R* ≤ *r*)/P(*R* ≤ *r*) = $\varphi\left( s \right)/\Phi\left( r \right)$, where $\varphi\left( s \right)$ is the probability density function of the standard normal distribution. We can gain an intuitive understanding of this result by setting *r* equal to zero (*i.e.* $\pi_{1}$ = 0.5), in which case $p(s)$ = $2\varphi\left( s \right)$. The records with observed values of *Y* will have values of *R* across the entire negative range of the standard normal distribution. By symmetry, each value of *R* is then twice as likely to occur in the complete records as in the full data.

If there are no missing values of *Y* (i.e. $\pi_{1}$ = 1), $p(s)$ equals $\varphi\left( s \right)$ and $\alpha_{1}^{OBS}$ is the average value of $\beta_{YX|W,R}$ across the full range of *R*, *i.e.* $\alpha_{1}^{OBS}=\int_{-\infty}^{\infty} \beta_{YX|W,R=s} \varphi\left( s \right)ds$, which is the marginal parameter, $\beta_{YX|W}$. Recall from the main text that $\beta_{YX|W}$ is equivalent to $\beta_{YX}$ in our scenario (because *X* is independent of *W* by construction). Therefore, as expected, the MI estimator $\beta_{YX}^{MI}$ is unbiased if all records are complete.

As the proportion of records with missing values of *Y* increases, we approach the situation in which $\pi_{1}$ = 0 (equivalent to the hypothetical scenario in which all values of *Y* are missing). We use a heuristic argument to evaluate $\alpha_{1}^{OBS}$ as $\pi_{1}$ tends to zero (or equivalently, since *r* = $\Phi^{-1}\left( \pi_{1} \right)$, as *r* tends to $-\infty$): consider a value of *r* such that the probability of observing a value less than *r* is approximately equal to 0 *i.e.* $\varphi\left( s \right)$ ≈ 0 for all *s* < *r*. In this case, $p(s)$ ≈ 0 for all *s* < *r*, with $p(r)$ ≈ 1. Then, (using summation to approximate integration) $\lim_{r\to-\infty} \alpha_{1}^{OBS}$ ≈ $\beta_{YX|W,R=r} p(r) +\sum_{s<r} \beta_{YX|W,R=s} p(s)$ = $\beta_{YX|W,R=r}$. Note that, since $\beta_{YX|W,R=r}$ has the same magnitude, regardless of the value of *r* (see Section S2), we use the more general expression, $\beta_{YX|W,R}$, hereafter. Therefore, we see that $\alpha_{1}^{OBS}$ tends to the conditional parameter $\beta_{YX|W,R}$ as $\pi_{1}$ tends to zero. Hence, in the hypothetical scenario in which all values of *Y* are missing, bias of the MI estimator takes its maximum value of $\left| \beta_{YX|W,R} - \beta_{YX} \right|$.

*Section S2. Derivation of the theoretical expression for the maximum bias of the MI estimator when a continuous outcome Y is partially observed and the imputation model includes collider variable W*

To derive an expression for the maximum bias, we need to derive an expression for $\beta_{YX|W,R}$ (since maximum bias = $\beta_{YX|W,R} - \beta_{YX}$). As per Figure 1 from the main text, we assume that *Y*, *X*, *Z*, *U*, *R*, and *W* are normally distributed, such that *Y* = $\beta_{YX}$*X* + $\beta_{YZ}$*Z* + $\varepsilon_{Y}$ where $\varepsilon_{Y}$ ~ N(0, $\sigma_{Y}^{2}$), *X* ~ N($\mu_{X}$, $\sigma_{X}^{2}$), *Z* ~ N($\mu_{Z}$, $\sigma_{Z}^{2}$), *U* ~ N($\mu_{U}$, $\sigma_{U}^{2}$), *R* = $\beta_{RX}$*X* + $\beta_{RU}$*U* + $\varepsilon_{R}$ where $\varepsilon_{R}$ ~ N(0, $\sigma_{R}^{2}$), and *W* = $\beta_{WZ}$*Z* + $\beta_{WU}$*U* + $\varepsilon_{W}$ where $\varepsilon_{W}$ ~ N(0, $\sigma_{W}^{2}$)

Then, using standard results, the joint distribution of *Y*, *X*, *Z*, *U*, *R*, and *W*, *f*(*Y*, *X*, *Z*, *U*, *R*, *W*), is multivariate normal with mean **µ**, covariance matrix **∑**, where:

**µ** = $\left( \begin{aligned} \beta_{YX} \mu_{X}+ \beta_{YZ} \mu_{Z} \\ \mu_{X} \\ \mu_{Z} \\ \mu_{U} \\ \beta_{RX} \mu_{X}+ \beta_{RU} \mu_{U} \\ \beta_{WZ} \mu_{Z} + \beta_{WU} \mu_{U} \end{aligned} \right)$

and **∑** =

$$\left( \begin{matrix} \beta_{YX}^{2}\sigma_{X}^{2}+{\beta_{YZ}^{2}\sigma}_{Z}^{2}+\sigma_{Y}^{2} & {\beta_{YX}\sigma}_{X}^{2} & \beta_{YZ}\sigma_{Z}^{2} & 0 & {\beta_{YX}\beta_{RX}\sigma}_{X}^{2} & {\beta_{YZ}\beta_{WZ}\sigma}_{Z}^{2} \\ {\beta_{YX}\sigma}_{X}^{2} & \sigma_{X}^{2} & 0 & 0 & {\beta_{RX}\sigma}_{X}^{2} & 0 \\ {\beta_{YZ}\sigma}_{Z}^{2} & 0 & \sigma_{Z}^{2} & 0 & 0 & {\beta_{WZ}\sigma}_{Z}^{2} \\ 0 & 0 & 0 & \sigma_{U}^{2} & {\beta_{RU}\sigma}_{U}^{2} & {\beta_{WU}\sigma}_{U}^{2} \\ {\beta_{YX}\beta_{RX}\sigma}_{X}^{2} & {\beta_{RX}\sigma}_{X}^{2} & 0 & {\beta_{RU}\sigma}_{U}^{2} & {\beta_{RX}^{2}\sigma}_{X}^{2}+{\beta_{RU}^{2}\sigma}_{U}^{2}+\sigma_{R}^{2} & {\beta_{RU}\beta_{WU}\sigma}_{U}^{2} \\ {\beta_{YZ}\beta_{WZ}\sigma}_{Z}^{2} & 0 & {\beta_{WZ}\sigma}_{Z}^{2} & {\beta_{WU}\sigma}_{U}^{2} & {\beta_{RU}\beta_{WU}\sigma}_{U}^{2} & {{\beta_{WZ}^{2}\sigma}_{Z}^{2}+\beta_{WU}^{2}\sigma}_{U}^{2}+\sigma_{W}^{2} \end{matrix} \right)$$

Then, again using standard results, the joint conditional distribution of *Y*, *X*, *Z,* and *U* given *R* and *W*, *f*(*Y*, *X*, *Z*, *U* | *R*= *r*, *W* = *w*), is multivariate normal with mean **µ***, covariance matrix **∑***, where:

**µ*** =$\left( \begin{aligned} \beta_{YX} \mu_{X}+ \beta_{YZ} \mu_{Z} \\ \mu_{X} \\ \mu_{Z} \\ \mu_{U} \end{aligned} \right)\boldsymbol{+}\left( \begin{matrix} {\beta_{YX}\beta_{RX}\sigma}_{X}^{2} & {\beta_{YZ}\beta_{WZ}\sigma}_{Z}^{2} \\ {\beta_{RX}\sigma}_{X}^{2} & 0 \\ 0 & {\beta_{WZ}\sigma}_{Z}^{2} \\ {\beta_{RU}\sigma}_{U}^{2} & {\beta_{WU}\sigma}_{U}^{2} \end{matrix} \right)\Sigma_{RW}^{-1}\left( \begin{matrix} r-\beta_{RX} \mu_{X}- \beta_{RU} \mu_{U} \\ w\boldsymbol{-}\beta_{WZ} \mu_{Z} - \beta_{WU} \mu_{U} \end{matrix} \right)$

where $\Sigma_{RW}^{-1}$ denotes the inverse of the covariance matrix for *R* and *W*, *i.e.* $\Sigma_{RW}^{-1}=\frac{1}{({\beta_{RX}^{2}\sigma}_{X}^{2}+{\beta_{RU}^{2}\sigma}_{U}^{2}+\sigma_{R}^{2})\left( {{\beta_{WZ}^{2}\sigma}_{Z}^{2}+\beta_{WU}^{2}\sigma}_{U}^{2}+\sigma_{W}^{2} \right)-{({\beta_{RU}\beta_{WU}\sigma}_{U}^{2})}^{2}}\left( \begin{matrix} {{\beta_{WZ}^{2}\sigma}_{Z}^{2}+\beta_{WU}^{2}\sigma}_{U}^{2}+\sigma_{W}^{2} & -{\beta_{RU}\beta_{WU}\sigma}_{U}^{2} \\ -{\beta_{RU}\beta_{WU}\sigma}_{U}^{2} & {\beta_{RX}^{2}\sigma}_{X}^{2}+{\beta_{RU}^{2}\sigma}_{U}^{2}+\sigma_{R}^{2} \end{matrix} \right)$

and

**∑*** = $\left( \begin{matrix} \beta_{YX}^{2}\sigma_{X}^{2}+{\beta_{YZ}^{2}\sigma}_{Z}^{2}+\sigma_{Y}^{2} & {\beta_{YX}\sigma}_{X}^{2} & \beta_{YZ}\sigma_{Z}^{2} & 0 \\ {\beta_{YX}\sigma}_{X}^{2} & \sigma_{X}^{2} & 0 & 0 \\ {\beta_{YZ}\sigma}_{Z}^{2} & 0 & \sigma_{Z}^{2} & 0 \\ 0 & 0 & 0 & \sigma_{U}^{2} \end{matrix} \right)-$

$\left( \begin{matrix} {\beta_{YX}\beta_{RX}\sigma}_{X}^{2} & {\beta_{YZ}\beta_{WZ}\sigma}_{Z}^{2} \\ {\beta_{RX}\sigma}_{X}^{2} & 0 \\ 0 & {\beta_{WZ}\sigma}_{Z}^{2} \\ {\beta_{RU}\sigma}_{U}^{2} & {\beta_{WU}\sigma}_{U}^{2} \end{matrix} \right)\Sigma_{RW}^{-1}\left( \begin{matrix} {\beta_{YX}\beta_{RX}\sigma}_{X}^{2} & {\beta_{RX}\sigma}_{X}^{2} & 0 & {\beta_{RU}\sigma}_{U}^{2} \\ {\beta_{YZ}\beta_{WZ}\sigma}_{Z}^{2} & 0 & {\beta_{WZ}\sigma}_{Z}^{2} & {\beta_{WU}\sigma}_{U}^{2} \end{matrix} \right)$

Expanding out the terms from **∑*** for Cov(*Y*, *X* | *W*, *R*) and Var(*X* | *W*, *R*) gives:

Cov(*Y*, *X* | *W*, *R*) = ${\beta_{YX}\sigma}_{X}^{2}-\frac{({{\beta_{WZ}^{2}\sigma}_{Z}^{2}+\beta_{WU}^{2}\sigma}_{U}^{2}+\sigma_{W}^{2})\beta_{YX}{\beta_{RX}^{2}\sigma}_{X}^{4}-\beta_{RX}\beta_{RU}\beta_{WU}\beta_{YZ}\beta_{WZ}\sigma_{Z}^{2}\sigma_{U}^{2}\sigma_{X}^{2}}{({\beta_{RX}^{2}\sigma}_{X}^{2}+{\beta_{RU}^{2}\sigma}_{U}^{2}+\sigma_{R}^{2})\left( {{\beta_{WZ}^{2}\sigma}_{Z}^{2}+\beta_{WU}^{2}\sigma}_{U}^{2}+\sigma_{W}^{2} \right)-{({\beta_{RU}\beta_{WU}\sigma}_{U}^{2})}^{2}}$

and

Var(*X* | *W*, *R*) = $\sigma_{X}^{2}-\frac{({{\beta_{WZ}^{2}\sigma}_{Z}^{2}+\beta_{WU}^{2}\sigma}_{U}^{2}+\sigma_{W}^{2}){\beta_{RX}^{2}\sigma}_{X}^{4}}{({\beta_{RX}^{2}\sigma}_{X}^{2}+{\beta_{RU}^{2}\sigma}_{U}^{2}+\sigma_{R}^{2})\left( {{\beta_{WZ}^{2}\sigma}_{Z}^{2}+\beta_{WU}^{2}\sigma}_{U}^{2}+\sigma_{W}^{2} \right)-{({\beta_{RU}\beta_{WU}\sigma}_{U}^{2})}^{2}}$

Hence, $\beta_{YX|W,R}$ = $\frac{\text{Cov(}\text{Y, X}\text{ | }\text{W, R}\text{)}}{\text{Var(}\text{X}\text{ | }\text{W, R}\text{)}}$ = $\left\{ \beta_{YX}\times\frac{\left( {\beta_{RX}^{2}\sigma}_{X}^{2}+{\beta_{RU}^{2}\sigma}_{U}^{2}+\sigma_{R}^{2} \right)\left( {{\beta_{WZ}^{2}\sigma}_{Z}^{2}+\beta_{WU}^{2}\sigma}_{U}^{2}+\sigma_{W}^{2} \right)-\left( {\beta_{RU}\beta_{WU}\sigma}_{U}^{2} \right)^{2}-\left( {{\beta_{WZ}^{2}\sigma}_{Z}^{2}+\beta_{WU}^{2}\sigma}_{U}^{2}+\sigma_{W}^{2} \right){\beta_{RX}^{2}\sigma}_{X}^{2}}{\left( {\beta_{RX}^{2}\sigma}_{X}^{2}+{\beta_{RU}^{2}\sigma}_{U}^{2}+\sigma_{R}^{2} \right)\left( {{\beta_{WZ}^{2}\sigma}_{Z}^{2}+\beta_{WU}^{2}\sigma}_{U}^{2}+\sigma_{W}^{2} \right)-\left( {\beta_{RU}\beta_{WU}\sigma}_{U}^{2} \right)^{2}-\left( {{\beta_{WZ}^{2}\sigma}_{Z}^{2}+\beta_{WU}^{2}\sigma}_{U}^{2}+\sigma_{W}^{2} \right){\beta_{RX}^{2}\sigma}_{X}^{2}} \right\}+\frac{\beta_{RX}\beta_{RU}\beta_{WU}\beta_{YZ}\beta_{WZ}\sigma_{Z}^{2}\sigma_{U}^{2}}{\left( {\beta_{RX}^{2}\sigma}_{X}^{2}+{\beta_{RU}^{2}\sigma}_{U}^{2}+\sigma_{R}^{2} \right)\left( {{\beta_{WZ}^{2}\sigma}_{Z}^{2}+\beta_{WU}^{2}\sigma}_{U}^{2}+\sigma_{W}^{2} \right)-\left( {\beta_{RU}\beta_{WU}\sigma}_{U}^{2} \right)^{2}-({{\beta_{WZ}^{2}\sigma}_{Z}^{2}+\beta_{WU}^{2}\sigma}_{U}^{2}+\sigma_{W}^{2}){\beta_{RX}^{2}\sigma}_{X}^{2}}$

= $\beta_{YX}+\frac{\beta_{RX}\beta_{RU}\beta_{WU}\beta_{YZ}\beta_{WZ}\sigma_{Z}^{2}\sigma_{U}^{2}}{\left( {\beta_{RU}^{2}\sigma}_{U}^{2}+\sigma_{R}^{2} \right)\left( {\beta_{WZ}^{2}\sigma}_{Z}^{2}+\sigma_{W}^{2} \right)+\beta_{WU}^{2}\sigma_{U}^{2}\sigma_{R}^{2}}$

Hence the maximum bias = $\beta_{YX|W,R} - \beta_{YX}$ = $\frac{\beta_{RX}\beta_{RU}\beta_{WU}\beta_{YZ}\beta_{WZ}\sigma_{Z}^{2}\sigma_{U}^{2}}{\left( {\beta_{RU}^{2}\sigma}_{U}^{2}+\sigma_{R}^{2} \right)\left( {\beta_{WZ}^{2}\sigma}_{Z}^{2}+\sigma_{W}^{2} \right)+\beta_{WU}^{2}\sigma_{U}^{2}\sigma_{R}^{2}}$ as per Formula 2.2.5 in the main text.

This can be written in terms of variance and covariance as:

$$\frac{\text{Cov}\left( \text{X}\text{,}\text{R} \right)\text{Cov}\left( \text{W}\text{,}\text{R} \right)\text{Cov}\left( \text{Y}\text{,}\text{W} \right)}{\left\{ \text{Var}\left( \text{R} \right)\text{Var}\text{(}\text{X}\text{)-}\text{Cov}^{\text{2}}\left( \text{X}\text{,}\text{R} \right) \right\}\text{Var}\left( \text{W} \right)\text{ - }\text{Var}\left( \text{X} \right)\text{Cov}^{\text{2}}\left( \text{W}\text{,}\text{R} \right)}$$

We note that, using similar derivations,

$\beta_{YX|W}$ = $\frac{\text{Cov}\text{(}\text{Y, X | W}\text{)}}{\text{Var}\text{(}\text{X | W}\text{)}}$ *=* $\frac{\text{Cov}\left( \text{Y, X} \right)\text{ - }\text{Cov}\left( \text{Y,W} \right)\text{Cov}\left( \text{X,W} \right)\text{/}\text{Var}\text{(}\text{W}\text{)}}{\text{Var}\text{(}\text{X}\text{)}\text{ -}\text{ }\text{Cov}^{\text{2}}\left( \text{X,W} \right)\text{/}\text{Var}\text{(}\text{W}\text{)}}$=$\beta_{YX}$ and $\beta_{YW|X}$=$\beta_{YW}$ since *Cov*(*X*,*W*) = 0

Similarly, $\beta_{YX|Z}$ = $\beta_{YX}$, $\beta_{YZ|R}$ = $\beta_{YZ}$, and $\beta_{YR|Z}$ = $\beta_{YR}$ since *Cov*(*X*,*Z*) = *Cov*(*Z*,*R*) = 0

and $\beta_{YX|R}$ = $\beta_{YX}$ since

$\beta_{YX|R}$ = $\frac{\text{Cov}\text{(}\text{Y, X |}\text{ }\text{R}\text{)}}{\text{Var}\text{(}\text{X | R}\text{)}}$ *=* $\frac{\text{Cov}\left( \text{Y, X} \right)\text{ - }\text{Cov}\left( \text{Y,R} \right)\text{Cov}\left( \text{X,R} \right)\text{/}\text{Var}\text{(}\text{R}\text{)}}{\text{Var}\text{(}\text{X}\text{)}\text{ -}\text{Cov}^{\text{2}}\left( \text{X,R} \right)\text{/}\text{Var}\text{(}\text{R}\text{)}}$ = $\beta_{YX}\times\frac{\text{Var(X)}\text{ - }\text{Cov}^{\text{2}}\left( \text{X,R} \right)\text{/}\text{Var}\text{(}\text{R}\text{)}}{\text{Var}\text{(}\text{X}\text{)}\text{ -}\text{Cov}^{\text{2}}\left( \text{X,R} \right)\text{/}\text{Var}\text{(}\text{R}\text{)}}$ = $\beta_{YX}$

*Section S3. Description of simulation study to illustrate the bias and standard error of the MI estimator when including a collider in the imputation model, compared with the standard error of the CRA estimator, as the proportion of missing data increases when a continuous outcome Y is partially observed.*

As per the main text, we illustrate how the bias and SE of the MI estimator when including a collider in the imputation model vary with $\pi_{0}$, using a simple simulation. For reference, we also illustrate how the SE of the CRA estimator varies with $\pi_{0}$ (the CRA estimator is always unbiased in this setting). This example is based on the relationships depicted in Figure 1 of the main text, setting the mean of each variable equal to zero, all direct effect sizes equal to one, and all error variances equal to one. Bias and SEs were estimated using 1000 simulated datasets per value of $\pi_{0}$ (with $\pi_{0}$ taking values in the range [0, 0.9]), with 100,000 records in each dataset. In addition, to illustrate the magnitude of bias as $\pi_{0}$ tends to one, bias was calculated for values of $\pi_{0}$ in the range [0.9,1), with the number of records increased to 10^6^ or 10^7^ observations to ensure Monte Carlo error for the complete records estimates was always sufficiently small. In each simulated dataset, a value of *Y* was set to missing if the corresponding value of *R* was greater than $\Phi^{-1}\left( 1-\pi_{0} \right)$. We used the fact that $\beta_{YX}^{MI}$ = $\alpha_{1}^{OBS}$ to estimate bias and SE of the MI estimator. Thus, for each value of $\pi_{0}$ (noting that, for brevity, in the following expressions we have suppressed the subscript which indicates that these are evaluated per value of $\pi_{0}$), bias was estimated as ${\bar{\hat{\alpha}}}_{1}^{OBS}-\beta_{YX}$, where $\beta_{YX}$ = 1 and ${\bar{\hat{\alpha}}}_{1}^{OBS}$ = $\sum_{i=1}^{1000} \frac{\hat{\alpha}_{1i}^{OBS}}{1000}$ denotes the mean of the per-simulation estimates of the *X* coefficient from a linear regression of *Y* on *X* conditional on *W*, fitted to the complete records *i.e.* those with observed values of *Y* (where $\hat{\alpha}_{1i}^{OBS}$ denotes the corresponding estimate for the *i^th^* simulated dataset); SEs of the MI and CRA estimators were calculated, assuming the number of records with an observed value of *Y* equals 1000, as: $\sqrt{\sum_{i=1}^{1000} \frac{{\hat{\text{SE}}}^{\text{2}}\left( \hat{\alpha}_{1i}^{OBS} \right)}{1000}}$🞨10$\sqrt{(1-\pi_{0})}$ and$\sqrt{\sum_{i=1}^{1000} \frac{{\hat{\text{SE}}}^{\text{2}}\left( \hat{\beta}_{YXi}^{OBS} \right)}{1000}}$🞨10$\sqrt{(1-\pi_{0})}$ ,respectively, where $\hat{\text{SE}}\left( \hat{\beta}_{YXi}^{OBS} \right)$ denotes the CRA estimate of $\beta_{YX}$ for the *i^th^* simulated dataset. We estimated $\beta_{YX}^{MI}$ by ${\bar{\hat{\alpha}}}_{1}^{OBS}$ rather than by the mean of the MI estimates of $\beta_{YX}$ (${\bar{\hat{\beta}}}_{YX}^{MI}$) to avoid an additional source of sampling error (because each per-simulation MI estimate is itself a pooled estimate of the per-imputation estimates). However, for $\pi_{0}$ in the range [0.1, 0.9], we verified empirically (using five imputations and 1000 observations in each dataset) that ${\bar{\hat{\beta}}}_{YX}^{MI}$ was unbiased for $\hat{\alpha}_{1}^{OBS}$.

*Figure S1. Estimated standard error (SE) of the MI estimator of* $\beta_{YX}$ *when the imputation model includes a collider, W, and SE of the complete records analysis (CRA) estimator of* $\beta_{YX}$*, plotted against the proportion of records with missing data, when continuous outcome Y is partially observed**, assuming 1000 observed values. The strength of the associations between both Y and Z, and W and Z equal 0.5, all other direct effect sizes and error variances equal one. Horizontal grey solid lines represent the values of SE of the MI estimator when the proportion of records with missing data is zero (lower line) or tends to one (upper line). Horizontal grey dashed lines represent the values of SE of the CRA estimator when the proportion of records with missing data is zero (lower line) or tends to one (upper line).*

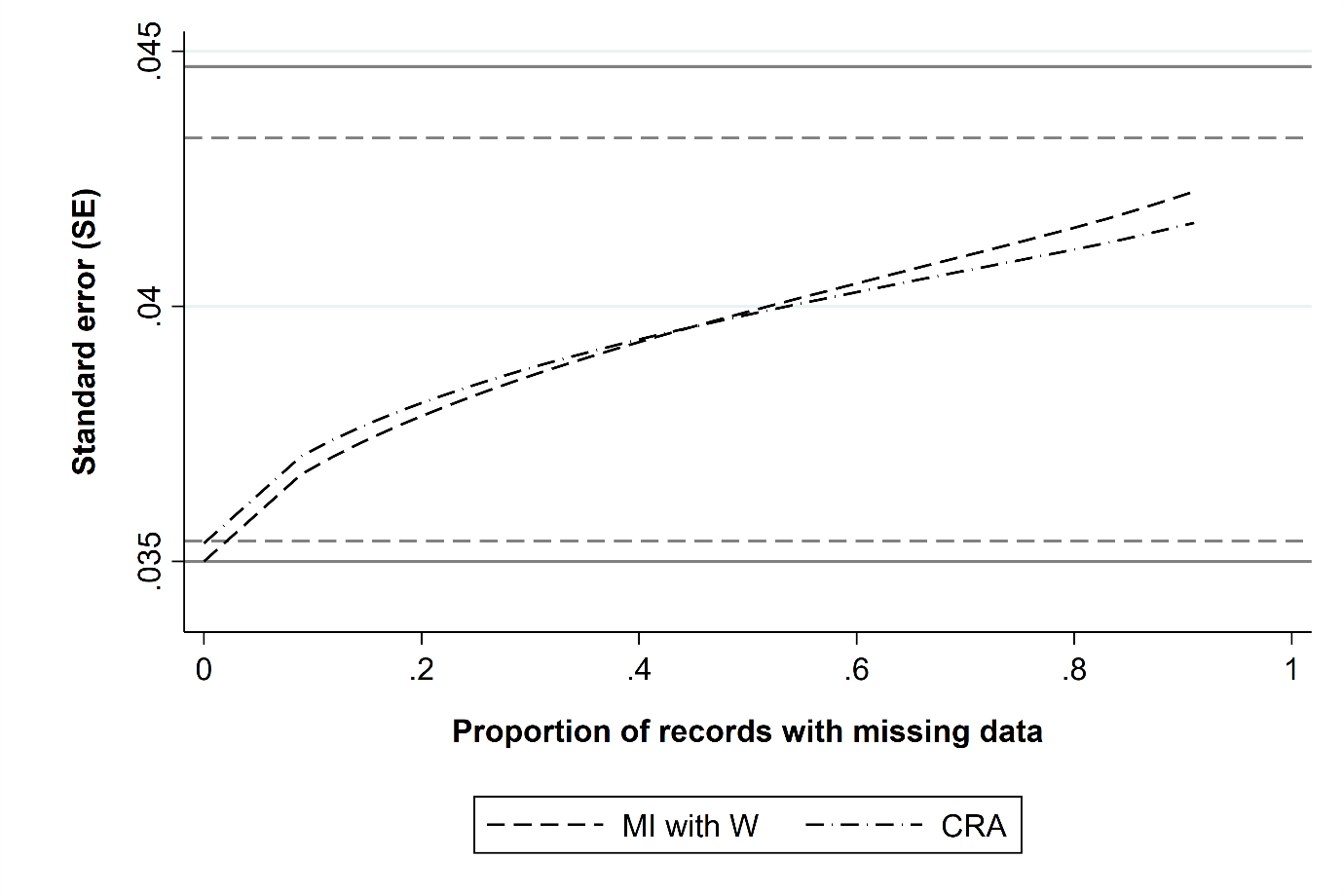

*Section S4. Verification of the theoretical expression for the maximum bias of the MI estimator when a continuous outcome Y is partially observed and the imputation model includes collider variable W*

The theoretical expression for the maximum bias of the MI estimator when *Y* is partially observed (and the imputation model includes *W*) was verified using simulation. We used 1000 simulations, and each simulated dataset contained 100,000 observations. In each simulated dataset, the values of each coefficient ($\beta_{YX}$, $\beta_{WZ}$, *etc*.) and each error variance ($\sigma_{X}^{2}$, $\sigma_{Z}^{2}$, *etc.*) were sampled from a uniform distribution *U*(0, 2). For simplicity, $\mu_{X}$, $\mu_{Z}$, and $\mu_{U}$ were set equal to zero (note that the maximum bias formula does not depend on these parameters). Data were then generated using the specified models for *Y*, *X*, *Z*, *U*, *R*, and *W* as per Section S2, above. Maximum bias was calculated using the theoretical expression (Formula 2.2.5 in the main text). Maximum bias was also estimated empirically by calculating the difference in the *X* coefficient when fitting a linear regression of (i) *Y* on *X*, conditional on *W* and *R*, and (ii) *Y* on *X*.

The median difference between the theoretical and empirical values of maximum bias was 0.000 (5^th^ – 95^th^ percentile: -0.016 - 0.013). Therefore, we conclude that the theoretical expression for maximum bias is correct.

*Section S5.*

*Figure S2. Relative increase in precision of the MI estimator of* $\beta_{YX}$ *when a collider is included in the imputation model compared with the CRA estimator, as the proportion of missing data tends to one. Results shown when continuous outcome Y is partially observed, varying direct effect sizes* $\beta_{YX}$*,* $\beta_{RX}$*,* $\beta_{RU}$*,* $\beta_{WU}$*,* $\beta_{YZ}$*, and* $\beta_{WZ}$*. The distribution of the relative increase in precision in each box-plot is averaged over the values of* $\beta_{RX}$, $\beta_{RU}$ *and* $\beta_{WU}$*.*

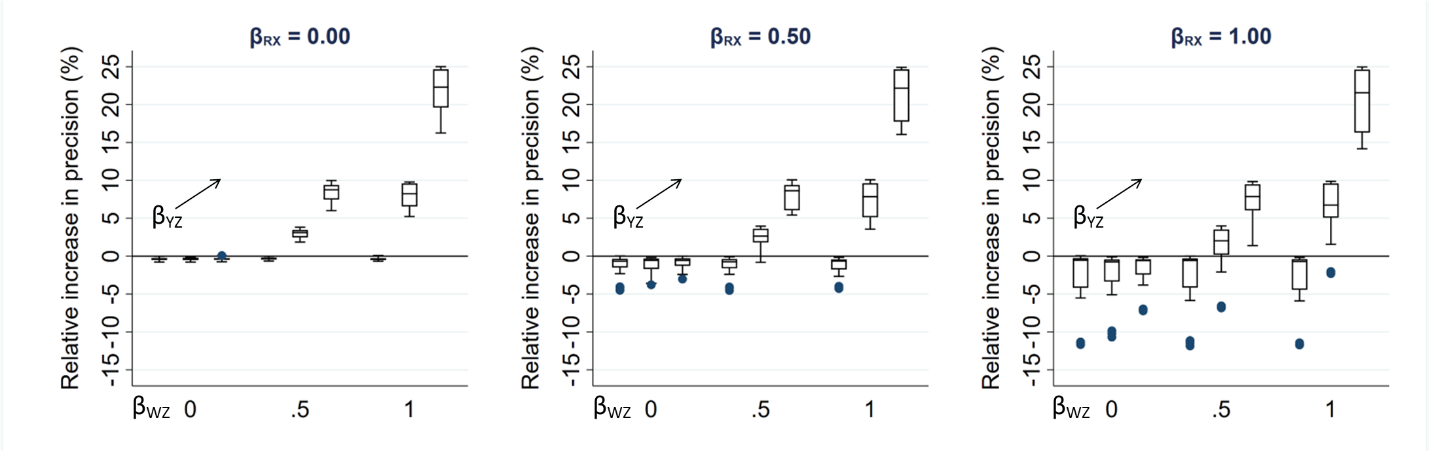

*Section S6. Derivation of the theoretical expression for the maximum bias of the Y coefficient in the imputation model for continuous exposure X when the imputation model includes collider variable W*

We start by briefly summarising the details from the main text: We derive an expression for the maximum bias of the *Y* coefficient in the imputation model for continuous exposure *X*, assuming that MI is performed by replacing missing values of *X* with draws from a linear regression model and hence, the imputation model for *X* is of the form: $\text{E}\left( \text{X} \right)\text{ = }\alpha_{0}\text{ + }\alpha_{1}\text{Y }\text{+ }\alpha_{2}\text{W}$. Then, the MI estimator of $\beta_{YX}$ will be unbiased only if $\alpha_{0}^{OBS}$ = $\alpha_{0}$, $\alpha_{1}^{OBS}$ = $\alpha_{1}$, and $\alpha_{2}^{OBS}$ = $\alpha_{2}$, where $\alpha_{.}^{OBS}$ denotes the parameter based on the records with observed values of *X*.

Taking $\alpha_{1}$ as an example, and using a similar argument to that in Section S1, $\alpha_{1}^{OBS} = \beta_{XY|W,R_{ind}=1} = \int_{-\infty}^{r} \beta_{XY|W,R=s} p(s)ds$ where $p(s)$ denotes the probability that *R* = *s* given *s* $\in(-\infty, \text{r}]$. When there are no missing values of *X*, $\alpha_{1}^{OBS}$ equals the marginal parameter, $\beta_{XY|W}$. In the hypothetical scenario in which all values of *X* are missing, $\alpha_{1}^{OBS}$ equals the conditional parameter $\beta_{XY|W,R}$ and the bias of $\alpha_{1}^{OBS}$ takes its maximum value of $\left| \beta_{XY|W,R} - \beta_{XY|W} \right|$.

As per Figure 4 from the main text, we assume that *Y*, *X*, *Z*, *W*, and *U* are normally distributed, and that there exists a normally distributed variable *R* such that P($\text{R}_{\text{ind}}$ = 1) = P(*R* ≤ r). We further assume that each of *Y*, *X*, *W*, and *R* is a linear combination of the variables causing it plus an error term (with *Z* and *U* having no direct causes), with no interactions, all errors uncorrelated, no model mis-specification, and no measurement error. Finally, we assume an ordinary least squares (OLS) estimator is used to obtain estimates in both analysis and imputation models.

Specifically, we define: *Y* = $\beta_{YX}$*X* + $\varepsilon_{Y}$ where $\varepsilon_{Y}$ ~ N(0, $\sigma_{Y}^{2}$), *X* = $\beta_{XZ}$*Z* + $\varepsilon_{X}$ where $\varepsilon_{X}$ ~ N(0, $\sigma_{X}^{2}$), *Z* ~ N($\mu_{Z}$, $\sigma_{Z}^{2}$), *U* ~ N($\mu_{U}$, $\sigma_{U}^{2}$), *R* = $\beta_{RU}$*U* + $\varepsilon_{R}$ where $\varepsilon_{R}$ ~ N(0, $\sigma_{R}^{2}$), and *W* = $\beta_{WZ}$*Z* + $\beta_{WU}$*U* + $\varepsilon_{W}$ where $\varepsilon_{W}$ ~ N(0, $\sigma_{W}^{2}$)

Then, using standard results, the joint distribution of *Y*, *X*, *Z*, *U*, *R*, and *W*, *f*(*Y*, *X*, *Z*, *U*, *R*, *W*), is multivariate normal with mean **µ**, covariance matrix **∑**, where:

**µ** = $\left( \begin{aligned} \beta_{YX} \beta_{XZ} \mu_{Z} \\ \beta_{XZ} \mu_{Z} \\ \mu_{Z} \\ \mu_{U} \\ \beta_{RU} \mu_{U} \\ \beta_{WZ} \mu_{Z} + \beta_{WU} \mu_{U} \end{aligned} \right)$

and **∑** =

$$\left( \begin{matrix} \beta_{YX}^{2}({\beta_{XZ}^{2}\sigma}_{Z}^{2}+\sigma_{X}^{2})+\sigma_{Y}^{2} & \beta_{YX}({\beta_{XZ}^{2}\sigma}_{Z}^{2}+\sigma_{X}^{2}) & \beta_{YX}\beta_{XZ}\sigma_{Z}^{2} & 0 & 0 & \beta_{YX}{\beta_{XZ}\beta_{WZ}\sigma}_{Z}^{2} \\ \beta_{YX}({\beta_{XZ}^{2}\sigma}_{Z}^{2}+\sigma_{X}^{2}) & {\beta_{XZ}^{2}\sigma}_{Z}^{2}+\sigma_{X}^{2} & {\beta_{XZ}\sigma}_{Z}^{2} & 0 & 0 & {\beta_{XZ}\beta_{WZ}\sigma}_{Z}^{2} \\ \beta_{YX}\beta_{XZ}\sigma_{Z}^{2} & {\beta_{XZ}\sigma}_{Z}^{2} & \sigma_{Z}^{2} & 0 & 0 & {\beta_{WZ}\sigma}_{Z}^{2} \\ 0 & 0 & 0 & \sigma_{U}^{2} & {\beta_{RU}\sigma}_{U}^{2} & {\beta_{WU}\sigma}_{U}^{2} \\ 0 & 0 & 0 & {\beta_{RU}\sigma}_{U}^{2} & {\beta_{RU}^{2}\sigma}_{U}^{2}+\sigma_{R}^{2} & {\beta_{RU}\beta_{WU}\sigma}_{U}^{2} \\ \beta_{YX}{\beta_{XZ}\beta_{WZ}\sigma}_{Z}^{2} & {\beta_{XZ}\beta_{WZ}\sigma}_{Z}^{2} & {\beta_{WZ}\sigma}_{Z}^{2} & {\beta_{WU}\sigma}_{U}^{2} & {\beta_{RU}\beta_{WU}\sigma}_{U}^{2} & {{\beta_{WZ}^{2}\sigma}_{Z}^{2}+\beta_{WU}^{2}\sigma}_{U}^{2}+\sigma_{W}^{2} \end{matrix} \right)$$

Then, again using standard results, the joint conditional distribution of *Y*, *X*, *Z,* and *U* given *R* and *W*, *f*(*Y*, *X*, *Z*, *U* | *R*= *r*, *W* = *w*), is multivariate normal with mean **µ***, covariance matrix **∑***, where:

**µ*** =$\left( \begin{aligned} \beta_{YX} \beta_{XZ} \mu_{Z} \\ \beta_{XZ} \mu_{Z} \\ \mu_{Z} \\ \mu_{U} \end{aligned} \right)\boldsymbol{+}\left( \begin{matrix} 0 & \beta_{YX}{\beta_{XZ}\beta_{WZ}\sigma}_{Z}^{2} \\ 0 & {\beta_{XZ}\beta_{WZ}\sigma}_{Z}^{2} \\ 0 & {\beta_{WZ}\sigma}_{Z}^{2} \\ {\beta_{RU}\sigma}_{U}^{2} & {\beta_{WU}\sigma}_{U}^{2} \end{matrix} \right)\Sigma_{RW}^{-1}\left( \begin{matrix} r-\beta_{RU} \mu_{U} \\ w\boldsymbol{-}\beta_{WZ} \mu_{Z} - \beta_{WU} \mu_{U} \end{matrix} \right)$

where $\Sigma_{RW}^{-1}$ denotes the inverse of the covariance matrix for *R* and *W*, *i.e.* $\Sigma_{RW}^{-1}=\frac{1}{({\beta_{RU}^{2}\sigma}_{U}^{2}+\sigma_{R}^{2})\left( {{\beta_{WZ}^{2}\sigma}_{Z}^{2}+\beta_{WU}^{2}\sigma}_{U}^{2}+\sigma_{W}^{2} \right)-{({\beta_{RU}\beta_{WU}\sigma}_{U}^{2})}^{2}}\left( \begin{matrix} {{\beta_{WZ}^{2}\sigma}_{Z}^{2}+\beta_{WU}^{2}\sigma}_{U}^{2}+\sigma_{W}^{2} & -{\beta_{RU}\beta_{WU}\sigma}_{U}^{2} \\ -{\beta_{RU}\beta_{WU}\sigma}_{U}^{2} & {\beta_{RU}^{2}\sigma}_{U}^{2}+\sigma_{R}^{2} \end{matrix} \right)$

and

**∑*** = $\left( \begin{matrix} \beta_{YX}^{2}({\beta_{XZ}^{2}\sigma}_{Z}^{2}+\sigma_{X}^{2})+\sigma_{Y}^{2} & \beta_{YX}({\beta_{XZ}^{2}\sigma}_{Z}^{2}+\sigma_{X}^{2}) & \beta_{YX}\beta_{XZ}\sigma_{Z}^{2} & 0 \\ \beta_{YX}({\beta_{XZ}^{2}\sigma}_{Z}^{2}+\sigma_{X}^{2}) & {\beta_{XZ}^{2}\sigma}_{Z}^{2}+\sigma_{X}^{2} & {\beta_{XZ}\sigma}_{Z}^{2} & 0 \\ \beta_{YX}\beta_{XZ}\sigma_{Z}^{2} & {\beta_{XZ}\sigma}_{Z}^{2} & \sigma_{Z}^{2} & 0 \\ 0 & 0 & 0 & \sigma_{U}^{2} \end{matrix} \right)-$

$\left( \begin{matrix} 0 & \beta_{YX}{\beta_{XZ}\beta_{WZ}\sigma}_{Z}^{2} \\ 0 & {\beta_{XZ}\beta_{WZ}\sigma}_{Z}^{2} \\ 0 & {\beta_{WZ}\sigma}_{Z}^{2} \\ {\beta_{RU}\sigma}_{U}^{2} & {\beta_{WU}\sigma}_{U}^{2} \end{matrix} \right)\Sigma_{RW}^{-1}\left( \begin{matrix} 0 & 0 & 0 & {\beta_{RU}\sigma}_{U}^{2} \\ \beta_{YX}{\beta_{XZ}\beta_{WZ}\sigma}_{Z}^{2} & {\beta_{XZ}\beta_{WZ}\sigma}_{Z}^{2} & {\beta_{WZ}\sigma}_{Z}^{2} & {\beta_{WU}\sigma}_{U}^{2} \end{matrix} \right)$

Expanding out the terms from **∑*** for Cov(*X, Y* | *W*, *R*) and Var(*Y* | *W*, *R*) gives:

Cov(*X*, *Y* |*W*, *R*) = $\beta_{YX}({\beta_{XZ}^{2}\sigma}_{Z}^{2}+\sigma_{X}^{2})-\frac{\beta_{YX}{(\beta}_{RU}^{2}\sigma_{U}^{2}+\sigma_{R}^{2})\left( {\beta_{XZ}\beta_{WZ}\sigma}_{Z}^{2} \right)^{2}}{({\beta_{RU}^{2}\sigma}_{U}^{2}+\sigma_{R}^{2})\left( {{\beta_{WZ}^{2}\sigma}_{Z}^{2}+\beta_{WU}^{2}\sigma}_{U}^{2}+\sigma_{W}^{2} \right)-{({\beta_{RU}\beta_{WU}\sigma}_{U}^{2})}^{2}}$

and

Var(*Y* |*W*, *R*) = $\beta_{YX}^{2}({\beta_{XZ}^{2}\sigma}_{Z}^{2}+\sigma_{X}^{2})+\sigma_{Y}^{2}-\frac{{(\beta}_{RU}^{2}\sigma_{U}^{2}+\sigma_{R}^{2})\left( \beta_{YX}{\beta_{XZ}\beta_{WZ}\sigma}_{Z}^{2} \right)^{2}}{({\beta_{RU}^{2}\sigma}_{U}^{2}+\sigma_{R}^{2})\left( {{\beta_{WZ}^{2}\sigma}_{Z}^{2}+\beta_{WU}^{2}\sigma}_{U}^{2}+\sigma_{W}^{2} \right)-{({\beta_{RU}\beta_{WU}\sigma}_{U}^{2})}^{2}}$

For brevity, we now proceed in terms of variance and covariance, expressing $\beta_{XY|W,R}$ as:

$\frac{\text{Cov(}\text{Y}\text{, }\text{X}\text{|}\text{W}\text{, }\text{R}\text{)}}{\text{Var(}\text{X}\text{|}\text{W}\text{, }\text{R}\text{)}}\text{=}\frac{\text{\{Var(}\text{R}\text{)Var(}\text{W}\text{) –} \text{Cov}^{\text{2}}\text{(}\text{R,W}\text{)\} Cov}\left( \text{Y,X} \right)\text{ - }\text{Var}\text{(}\text{R}\text{)}\text{Cov}\text{(}\text{Y}\text{,}\text{W}\text{)}\text{Cov}\text{(}\text{X}\text{,}\text{W}\text{)}}{\text{\{Var(}\text{R}\text{)Var(}\text{W}\text{) –} \text{Cov}^{\text{2}}\text{(}\text{R,W}\text{)\}Var}\left( \text{Y} \right)\text{ - }\text{Var}\text{(}\text{R}\text{)}\text{Cov}^{\text{2}}\text{(}\text{Y}\text{,}\text{W}\text{)}}$

Using a similar derivation, $\beta_{XY|W}$ = $\frac{\text{Cov}\text{(}\text{Y,X|W}\text{)}}{\text{Var(}\text{Y|W}\text{)}}$ = $\frac{\text{Cov}\left( \text{Y,X} \right)\text{- }\frac{\text{Cov}\left( \text{Y}\text{,}\text{W} \right)\text{Cov}\left( \text{X}\text{,}\text{W} \right)}{\text{Var(}\text{W}\text{)}}}{\text{Var}\left( \text{Y} \right)\text{ - }\frac{\text{Cov}^{\text{2}}\left( \text{Y}\text{,}\text{W} \right)}{\text{Var(}\text{W}\text{)}}}$, which, in passing, we note is not equal to $\beta_{XY}$ (because *X* and *Y* are not independent of *W*).

Hence, $\beta_{XY|W,R}$ can be expressed as $\frac{\text{Cov}\left( \text{Y,X|W} \right)\text{ – ACov(}\text{Y,X}\text{)}}{\text{Var}\left( \text{Y|W} \right)\text{ – AVar(}\text{Y}\text{)}}$ where A = $\frac{\text{Cov}^{\text{2}}\left( \text{R,W} \right)}{\text{Var(}\text{R}\text{)Var(}\text{W}\text{)}}$

Hence, the maximum bias of $\alpha_{1}^{OBS}$ = $\beta_{XY|W, R}- \beta_{XY|W}$

=$\frac{\text{Var}\left( \text{Y|W} \right)\left\{ \text{Cov}\left( \text{Y,X|W} \right)\text{ – ACov}\left( \text{Y,X} \right) \right\}\text{ – Cov}\left( \text{Y,X|W} \right)\text{\{Var}\left( \text{Y|W} \right)\text{ – AVar}\left( \text{Y} \right)\text{\} }}{\text{Var}\left( \text{Y|W} \right)\text{\{Var}\left( \text{Y|W} \right)\text{ – AVar}\left( \text{Y} \right)\text{\}}}$

=$\frac{\text{ A\{Var}\left( \text{Y} \right)\text{Cov}\left( \text{Y,X|W} \right)\text{ – }\text{Cov}\left( \text{Y}\text{,}\text{X} \right)\text{Var}\left( \text{Y|W} \right)\text{\} }}{\text{Var}\left( \text{Y|W} \right)\text{\{Var}\left( \text{Y|W} \right)\text{ – AVar}\left( \text{Y} \right)\text{\}}}$ as per the main text.

Using the same approach as in Section S4, the theoretical expression for the maximum bias of $\alpha_{1}^{OBS}$ was verified using simulation. We used 1000 simulations, and each simulated dataset contained 100,000 observations. In each simulated dataset, the values of each coefficient ($\beta_{YX}$, $\beta_{WZ}$, *etc*.) and each error variance ($\sigma_{X}^{2}$, $\sigma_{Z}^{2}$, *etc.*) were sampled from a uniform distribution *U*(0, 2). For simplicity, $\mu_{X}$, $\mu_{Z}$, and $\mu_{U}$ were set equal to zero (note that the maximum bias formula does not depend on these parameters). Data were then generated using the specified models for *Y*, *X*, *Z*, *U*, *R*, and *W* as above. Maximum bias was calculated using the theoretical expression above. Maximum bias was also estimated empirically by calculating the difference in the *Y* coefficient when fitting a linear regression of (i) *X* on *Y*, conditional on *W* and *R*, and (ii) *X* on *Y*, conditional on *W*.

The median difference between the theoretical and empirical values of maximum bias was 0.0000 (5^th^ – 95^th^ percentile: -0.0004 - 0.0005). Therefore, we conclude that the theoretical expression for maximum bias of $\alpha_{1}^{OBS}$ is correct.

*Section S7. Description of simulation study to assess bias of the MI estimator (and also relative increase in precision in the first setting) from including a collider in the imputation model when (i) continuous exposure X, (ii) binary outcome Y, or (iii) binary exposure X is partially observed.*

We performed simulation studies to assess bias of the MI estimator from including a collider in the imputation model when 50% of values were missing for a (i) continuous exposure X, (ii) binary outcome Y, or (iii) binary exposure X. In setting (i), we also assessed the relative precision of the MI estimator compared with the CRA estimator. In each setting, 1000 simulated datasets of size 1000 were generated using the data generating mechanisms described below. We used moderate values of the direct effect sizes (relative to the error variances, which were all equal to one), with $\beta_{YX}$ set to 0.00, 0.50, or 1.00 and all other direct effect sizes set to 0.00, 0.50, or 1.00 in setting (i) and 0.50 or 1.00 in settings (ii) and (iii). We assumed each variable had a mean of zero. We then set 50% of values of the partially observed variable to missing (in each case, by setting values to missing if *R* > 0). Bias in the MI estimate of $\beta_{YX}$ was calculated as the average of the per-simulation bias in each case (using W as an auxiliary variable in an imputation model that also included the completely observed analysis model variable, with five imputations). In setting (i), SE of the MI and CRA estimates were also calculated as the square root of the mean of the per-simulation squared SEs, and the relative increase in precision was calculated as 100 🞨(1 - (SE of the MI estimate)^2^/(SE of the CRA estimate)^2^).

*Data generating mechanisms*

In settings (i) and (iii) (continuous or binary exposure *X* is partially observed), in which *Y* was not a cause of missingness in *X*, continuous variables *Y*, *Z*, *U*, *R*, and *W* were related as defined in Section S6, with *X* defined as in Section S6 when *X* was continuous, and *X* defined as *logit*{P(*X*=1)} *=* $\beta_{XZ}$*Z* when *X* was binary. In the additional version of setting (i) in which *Y* was a cause of missingness in *X*, variables *X*, *Y*, *Z*, *U*, and *W* were related as defined in Section S6, and *R* was defined as *R* = $\beta_{RY}$*Y* + $\beta_{RU}$*U* + $\varepsilon_{R}$ where $\varepsilon_{R}$ ~ N(0, $\sigma_{R}^{2}$). In setting (ii) (binary outcome *Y* is partially observed), continuous variables *X*, *Z*, *U*, *R*, and *W* were related as defined in Section S2. *Y* was a binary variable such that *logit*{P(*Y*=1)} *=* $\beta_{YX}$*X* + $\beta_{YZ}$*Z*.

*Results*

Relative increase in precision of the MI estimate of $\beta_{YX}$ when 50% of values of a continuous exposure *X* are missing, and bias of the MI estimate of $\beta_{YX}$ when 50% of values of a binary outcome *Y* or binary exposure *X* are missing, are illustrated in Figures S3 to S5, respectively.

*Figure S3. Relative increase in precision of the MI estimate of* $\beta_{YX}$ *when a collider is included in the imputation model, compared with the CRA estimate. Results shown when 50% of values of a continuous exposure X are missing, varying direct effect sizes* $\beta_{YX}$*,* $\beta_{XZ}$*,* $\beta_{WZ}$*,* $\beta_{RU}$*, and* $\beta_{WU}$*. The distribution of the relative increase in precision in each box-plot is averaged over the values of* $\beta_{RU}$ *and* $\beta_{WU}$*.*

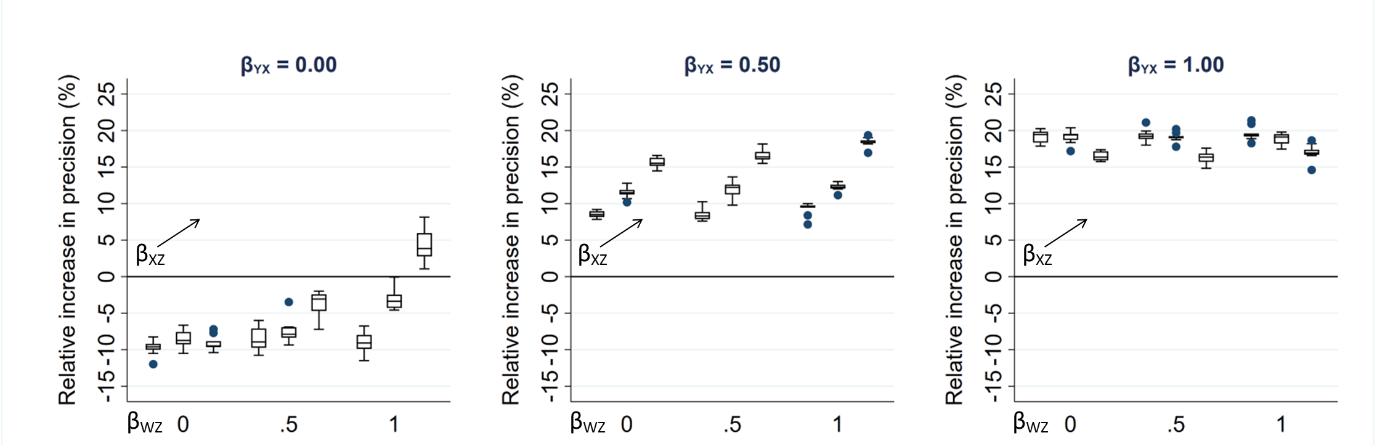

*Figure S4. Bias of the MI estimate of* $\beta_{YX}$ *when 50% of values of a binary outcome Y are missing, varying direct effect sizes* $\beta_{YX}$*,* $\beta_{RX}$*,* $\beta_{RU}$*,* $\beta_{WU}$*,* $\beta_{YZ}$*, and* $\beta_{WZ}$*. The distribution of maximum bias in each box-plot is averaged over the values of* $\beta_{RX}$, $\beta_{RU}$ *and* $\beta_{WU}$*.*

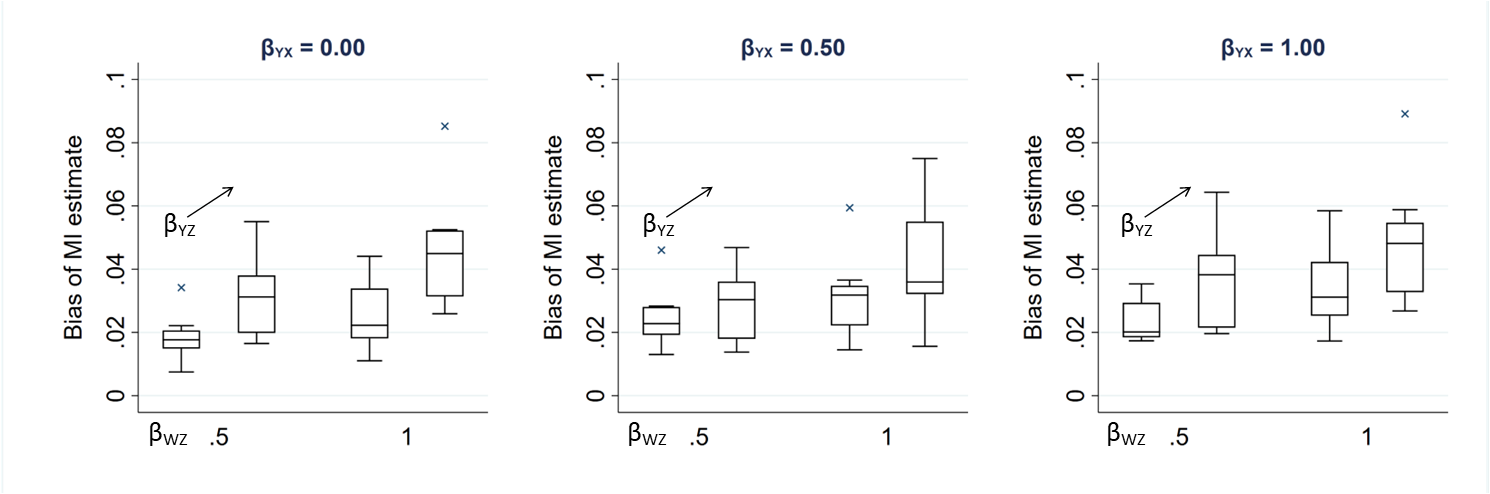

*Figure S5. Bias of the MI estimate of* $\beta_{YX}$ *when 50% of values of a binary exposure X are missing, varying direct effect sizes* $\beta_{YX}$*,* $\beta_{XZ}$*,* $\beta_{WZ}$*,* $\beta_{RU}$*, and* $\beta_{WU}$*. The distribution of bias in each box-plot is averaged over the values of* $\beta_{RU}$ *and* $\beta_{WU}$*.*

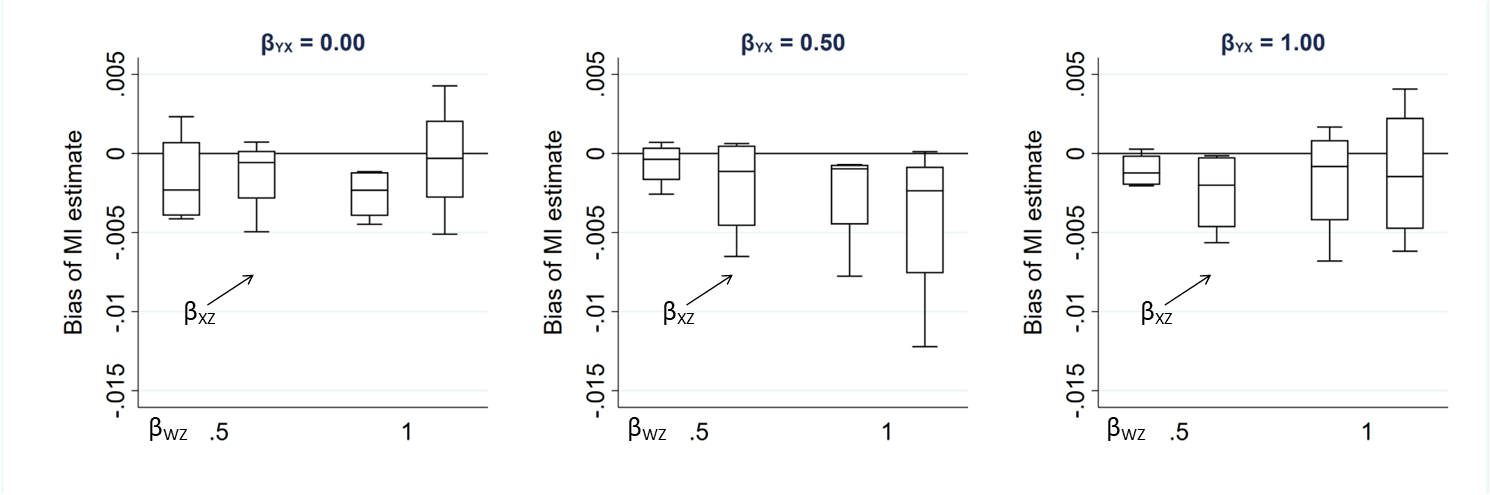

*Section S8. Stata code for formula verification and illustration, and to generate data as per the simulation studies*

```
/*1. Illustration of the bias and standard error of the MI estimator when including a collider in the imputation model when continuous outcome Y is partially observed */
clear
set seed 62543
*Define postfile to store results
tempname simloop
postfile `simloop' int(i j) float(comp_recs beta_xy_cond_w beta_xy_cond_w_SE beta_xy beta_xy_SE) using "sim_comp_case_beta_xy_cond_w_scen2.dta", replace

*Create a temporary file for storing full data
tempfile tmpfull

forvalues i=1/1000 {
	di "`i'"
	clear
	quietly set obs 100000
	gen z=rnormal(0,1)
	gen u=rnormal(0,1)
	gen x=rnormal(0,1)
	gen r=rnormal(x+u,1)
	gen w=rnormal(z+u,1)
	gen y=rnormal(x+z,1)

	quietly save `tmpfull', replace

	forvalues j=0/99 {
		use `tmpfull', clear
		gen ymiss=y
		quietly replace ymiss=. if r>invnormal(`j'/100)

		quietly regress ymiss x w
		local comp_recs = e(N)
		local beta_xy_cond_w = e(b)[1,1]
		local beta_xy_cond_w_SE =sqrt(e(V)[1,1])

		quietly regress ymiss x

post `simloop' (`i') (`j') (`comp_recs') (`beta_xy_cond_w')  (`beta_xy_cond_w_SE') (e(b)[1,1]) (sqrt(e(V)[1,1]))
	}
}
postclose `simloop'
/* Similar code used to add more granular measures at the extremes, not included to avoid repetition */

*Add MI estimates (with much smaller sample size as very slow to run)
clear
set seed 62543

*Define postfile to store results
tempname simloop
postfile `simloop' int(i j) float(comp_recs beta_x se_x) using "sim_MI_beta_xy_cond_w_scen2100000.dta", replace

*Create a temporary file for storing full data
tempfile tmpfull

forvalues i=1/1000 {
	di "`i'"
	clear
	quietly set obs 100000
	gen z=rnormal(0,1)
	gen u=rnormal(0,1)
	gen x=rnormal(0,1)
	gen r=rnormal(x+u,1)
	gen w=rnormal(z+u,1)
	gen y=rnormal(x+z,1)

	quietly save `tmpfull', replace

	forvalues j=1/99 {
		use `tmpfull', clear
		gen ymiss=y
		quietly replace ymiss=. if r>invnormal(`j'/100)

		quietly mi set flong
		quietly mi register imputed ymiss
		quietly mi register regular x w
		quietly mi impute chained (regress) ymiss = x w, add(5)
		local comp_recs=r(N_complete)[1,1]
		quietly mi estimate: regress ymiss x

post `simloop' (`i') (`j') (`comp_recs') (e(b_mi)[1,1]) (e(V_mi)[1,1])
	}
}
postclose `simloop'
postclose `simseed'

/*2. Verification of the maximum bias formula */
clear
set seed 5336665

*Define postfile to store results
tempname simloop
postfile `simloop' int(i) float(b_xy beta_xy beta_xy_cond_wr theor_bias emp_bias) using "sim_yxuvwr_scen2v3.dta", replace

forvalues i=1/1000 {
	clear
	*SD
	local s_z=runiform(0,2)
	local s_u=runiform(0,2)
	local s_r=runiform(0,2)
	local s_w=runiform(0,2)
	local s_x=runiform(0,2)
	local s_y=runiform(0,2)
	*beta
	local b_ur=runiform(0,2)
	local b_xr=runiform(0,2)
	local b_zw=runiform(0,2)
	local b_uw=runiform(0,2)
	local b_zy=runiform(0,2)
	local b_xy=runiform(0,2)

	*RVs
	quietly set obs 100000
	gen z=rnormal(0,`s_z')
	gen u=rnormal(0,`s_u')
	gen x=rnormal(0,`s_x')
	gen r=rnormal(`b_xr'*x + `b_ur'*u,`s_r')
	gen w=rnormal(`b_zw'*z+`b_uw'*u,`s_w')
	gen y=rnormal(`b_xy'*x + `b_zy'*z,`s_y')
	gen rind=(r<0)

	quietly regress y x
	local beta_xy=e(b)[1,1]

	*Estimate beta_XY|R,W
	quietly regress y x r w
	local beta_xy_cond_wr=e(b)[1,1]

	*Calculate theoretical and empirical bias
	local emp_bias = `beta_xy_cond_wr' - `beta_xy'

	local theor_bias = (`b_xr'*`b_ur'*`b_uw'*`b_zy'*`b_zw'*`s_z'^2*`s_u'^2)/((`s_r'^2 + `b_ur'^2*`s_u'^2)*(`b_zw'^2*`s_z'^2 + `s_w'^2 )+(`b_uw'^2*`s_u'^2*`s_r'^2))

	post `simloop' (`i') (`b_xy') (`beta_xy') (`beta_xy_cond_wr') (`theor_bias') (`emp_bias')
}
postclose `simloop'

/*3. Illustration of the maximum bias formula */
* Varying the size of parameters - still use mu_z=0 and mu_u=0 and all error vars = 1
set seed 62543

*Define postfile to store results
tempname simloop
postfile `simloop' float(i j k l m max_bias) using "maxbias_illustration_scen2.dta", replace

foreach i of numlist 0 0.25 0.5 0.75 1 {
	foreach j of numlist 0 0.25 0.5 0.75 1 {
		foreach k of numlist 0 0.25 0.5 0.75 1 {
			foreach l of numlist 0 0.25 0.5 0.75 1 {
				foreach m of numlist 0 0.25 0.5 0.75 1 {
					clear
					local b_rx=`i'
					local b_ru=`j'
					local b_wu=`k'
					local b_yz=`l'
					local b_wz=`m'


local max_bias=(`b_rx'*`b_ru'*`b_wu'*`b_yz'*`b_wz')/((1 + `b_ru'^2)*(1 + `b_wz'^2)+(`b_wu'^2))

post `simloop' (`i') (`j') (`k') (`l') (`m') (`max_bias')

				}
			}
		}
	}
}
postclose `simloop'

/* 4. Illustration of SE when max bias occurs */
*Varying the size of parameters - still use mu_z=0 and mu_u=0 and all error vars = 1
set seed 62543

*Define postfile to store results
tempname simloop
postfile `simloop' float(nsim b_rx b_ru b_wu b_yz b_wz b_yx beta_MI_SE) using "maxbias_illustration_scen2_SE.dta", replace

forvalues nsim=1/1000 {
	di "`nsim'"
	 foreach i of numlist 0 0.5 1 {
		foreach j of numlist 0 0.5 1 {
			foreach k of numlist 0 0.5 1 {
				foreach l of numlist 0 0.5 1 {
					foreach m of numlist 0 0.5 1 {
						foreach n of numlist 0 0.5 1 {
							clear
							quietly set obs 1000
							gen z=rnormal(0,1)
							gen u=rnormal(0,1)
							gen x=rnormal(0,1)
							gen r=rnormal(`i'*x + `j'*u,1)
							gen w=rnormal(`m'*z+`k'*u,1)
							gen y=rnormal(`n'*x + `l'*z,1)
							quietly regress y x w
post `simloop' (`nsim') (`i') (`j') (`k') (`l') (`m') (`n') (_se[x])						}
					}
				}
			}
		}
	}
}
postclose `simloop'

/* 5. Verification of the maximum bias formula for the Y coefficient in the imputation model for X when this also includes W  */
set seed 5336665

*Define postfile to store results
tempname simloop
postfile `simloop' int(i) float(beta_xy_cond_w theor_b_xy_cond_w beta_xy_cond_rw theor_bias emp_bias) using "sim_yxuvwr_scen1.dta", replace
forvalues i=1/1000 {
	clear
	*SD
	local s_z=runiform(0,2)
	local s_u=runiform(0,2)
	local s_r=runiform(0,2)
	local s_w=runiform(0,2)
	local s_x=runiform(0,2)
	local s_y=runiform(0,2)
	*beta
	local b_ur=runiform(0,2)
	local b_zw=runiform(0,2)
	local b_uw=runiform(0,2)
	local b_zx=runiform(0,2)
	local b_xy=runiform(0,2)
	*RVs
	quietly set obs 100000
	gen z=rnormal(0,`s_z')
	gen u=rnormal(0,`s_u')
	gen r=rnormal(`b_ur'*u,`s_r')
	gen w=rnormal(`b_zw'*z+`b_uw'*u,`s_w')
	gen x=rnormal(`b_zx'*z,`s_x')
	gen y=rnormal(`b_xy'*x,`s_y')

	*Estimate beta_XY|W
	quietly regress x y w
	local beta_xy_cond_w=e(b)[1,1]

	*Also store theor value
	local num = (`b_zw'^2*`s_z'^2 + `b_uw'^2*`s_u'^2 + `s_w'^2)*(`b_xy'*(`b_zx'^2*`s_z'^2 + `s_x'^2)) - `b_xy'*`b_zw'^2*`b_zx'^2*`s_z'^4
	local denom = (`b_zw'^2*`s_z'^2 + `b_uw'^2*`s_u'^2 + `s_w'^2)*(`b_xy'^2*(`b_zx'^2*`s_z'^2 + `s_x'^2) + `s_y'^2) - (`b_xy'^2*`b_zw'^2*`b_zx'^2*`s_z'^4)
	local theor_b_xy_cond_w = `num'/`denom'

	*local diff_beta_xy=e(b)[1,1] - (`b_xy'*(`b_zx'^2*`s_z'^2 + `s_x'^2))/(`b_xy'^2*(`b_zx'^2*`s_z'^2 + `s_x'^2) + `s_y'^2)

	*Estimate beta_XY|R,W
	quietly regress x y r w

	*Calculate theoretical and empirical bias
	local emp_bias = e(b)[1,1] - `beta_xy_cond_w'

	*Compare with calculated value
	local a=(`s_r'^2 + `b_ur'^2*`s_u'^2)*(`b_zw'^2*`s_z'^2 + `s_w'^2) + (`b_uw'^2*`s_r'^2*`s_u'^2)
	local b=(`s_r'^2 + `b_ur'^2*`s_u'^2)*`b_xy'*`b_zw'^2*`b_zx'^2*`s_z'^4
	local theor_bias=((`a'*(`b_xy'*(`b_zx'^2*`s_z'^2 + `s_x'^2)) - `b')/(`a'*(`b_xy'^2*(`b_zx'^2*`s_z'^2 + `s_x'^2) + `s_y'^2) - `b_xy'*`b')) - `theor_b_xy_cond_w'

post `simloop' (`i')  (`beta_xy_cond_w')  (`theor_b_xy_cond_w')  (e(b)[1,1])  (`theor_bias')  (`emp_bias')
}
postclose `simloop'

/* 6. Data generation when 50% of values of a continuous exposure X are missing */
* Varying the size of parameters - still use mu_z=0 and mu_u=0 and all error vars = 1
set seed 62543

tempname simloop
postfile `simloop' int(nsim) float(b_yx b_ru b_wu b_xz b_wz beta_xMI se_xMI beta_xCRA se_xCRA) using "MIbias_illustration_scen1Xcts.dta", replace

forvalues nsim=1/1000 {
	di "`nsim'"
	foreach i of numlist 0 0.5 1 {
		foreach j of numlist 0 0.5 1 {
			foreach k of numlist 0 0.5 1 {
				foreach l of numlist 0 0.5 1 {
					foreach m of numlist 0 0.5 1 {
					clear
					quietly set obs 1000
					*local b_yx=`i'
					*local b_ru=`j'
					*local b_wu=`k'
					*local b_xz=`l'
					*local b_wz=`m'

					gen z=rnormal(0,1)
					gen u=rnormal(0,1)
					gen r=rnormal(`j'*u,1)
					gen w=rnormal(`m'*z+`k'*u,1)
					gen x=rnormal(`l'*z,1)
					gen y=rnormal(`i'*x,1)

					gen xmiss=x
					quietly replace xmiss=. if r>0

*CRA
quietly regress y xmiss
local beta_xCRA= e(b)[1,1]
local se_xCRA= sqrt(e(V)[1,1])

		*MI with W
		quietly mi set flong
		quietly mi register imputed xmiss
		quietly mi register regular y w
		quietly mi impute chained (regress) xmiss = y w, add(5)
		quietly mi estimate: regress y xmiss

post `simloop' (`nsim') (`i') (`j') (`k') (`l') (`m') (e(b_mi)[1,1]) (sqrt(e(V_mi)[1,1])) (`beta_xCRA’) (`se_xCRA’)
					}
				}
			}
		}
	}
}
postclose `simloop'

/* 7. Data generation when 50% of values of a binary outcome Y are missing */
* Varying the size of parameters - still use mu_z=0 and mu_u=0 and all error vars = 1
set seed 62543

tempname simloop
postfile `simloop' int(nsim) float(b_yx b_rx b_ru b_wz b_uw b_yz beta_x se_x) using "MIbias_illustration_scen2Ybin.dta", replace
forvalues nsim=1/1000 {
	di "`nsim'"
	foreach i of numlist 0 0.5 1 {
		foreach j of numlist 0.5 1 {
			foreach k of numlist 0.5 1 {
				foreach l of numlist 0.5 1 {
					foreach m of numlist 0.5 1 {
						foreach n of numlist 0.5 1 {
				clear
				quietly set obs 1000

				gen z=rnormal(0,1)
				gen u=rnormal(0,1)
				gen x=rnormal(0,1)
				gen r=rnormal(`j'*x + `k'*u,1)
				gen w=rnormal(`l'*z+`m'*u,1)
				gen y=rbinomial(1,invlogit(`i'*x + `n'*z))

				gen ymiss=y
				quietly replace ymiss=. if r>0

				*MI with W
				quietly mi set flong
				quietly mi register imputed ymiss
				quietly mi register regular x w
				quietly mi impute chained (logit) ymiss = x w, add(5)
				quietly mi estimate: logistic ymiss x

	post `simloop' (`nsim') (`i') (`j') (`k') (`l') (`m') (`n') (e(b_mi)[1,1]) (sqrt(e(V_mi)[1,1]))
						}
					}
				}
			}
		}
	}
}
postclose `simloop'

/* 8. Data generation when 50% of values of a binary exposure X are missing */
set seed 62543

tempname simloop
postfile `simloop' int(nsim) float(b_yx b_ru b_wu b_xz b_wz beta_x se_x) using "MIbias_illustration_scen1Xbin.dta", replace

forvalues nsim=1/1000 {
	di "`nsim'"

	foreach i of numlist 0 0.5 1 {
		foreach j of numlist 0.5 1 {
			foreach k of numlist 0.5 1 {
				foreach l of numlist 0.5 1 {
					foreach m of numlist 0.5 1 {
		clear
		quietly set obs 1000
		gen z=rnormal(0,1)
		gen u=rnormal(0,1)
		gen r=rnormal(`j'*u,1)
		gen w=rnormal(`m'*z+`k'*u,1)
		gen x=rbinomial(1,invlogit(`l'*z))
		gen y=rnormal(`i'*x,1)

		gen xmiss=x
		quietly replace xmiss=. if r>0

		*MI with W
		quietly mi set flong
		quietly mi register imputed xmiss
		quietly mi register regular y w
quietly mi impute chained (logit) xmiss = y w, add(5)
		quietly mi estimate: regress y xmiss

		post `simloop' (`nsim') (`i') (`j') (`k') (`l') (`m') (e(b_mi)[1,1]) (sqrt(e(V_mi)[1,1]))
					}
				}
			}
		}
	}
}
postclose `simloop'
```

*Section S9. Stata code to perform the real data analysis*

*CRA of exposure outcome relationship

use alspac_subset, clear

regress bmi7 i.mated_binary

*MI linear imputation model with no aux variables

use alspac_subset, clear

quietly mi set flong

quietly mi register imputed bmi7

quietly mi register regular mated_binary

mi impute chained (regress) bmi7 = i.mated_binary, add(100) dots

mi estimate: regress bmi7 i.mated_binary

*MI linear imputation model including W=bwt

use alspac_subset, clear

quietly mi set flong

quietly mi register imputed bmi7

quietly mi register regular mated_binary bwt

mi impute chained (regress) bmi7 = i.mated_binary bwt, add(100) dots

mi estimate: regress bmi7 i.mated_binary

* Repeat for Z=pregsize_n

use alspac_subset, clear

quietly mi set flong

quietly mi register imputed bmi7

quietly mi register regular mated_binary pregsize

mi impute chained (regress) bmi7 = i.mated_binary i.pregsize, add(100) dots

mi estimate: regress bmi7 i.mated_binary

* And if use both pregsize_n and bwt

use alspac_subset, clear

quietly mi set flong

quietly mi register imputed bmi7

quietly mi register regular mated_binary pregsize bwt

mi impute chained (regress) bmi7 = i.mated_binary i.pregsize bwt, add(100) dots

mi estimate: regress bmi7 i.mated_binary

**** Theor max bias ******

use alspac_subset, clear

gen bwt_kg=bwt/1000

corr bmi7 bwt_kg, covariance

gen var_W=r(Var_2)

gen cov_YW=r(cov_12)

tab1 mated_binary

sum mated_binary

gen var_X=4256*7805/12061^2

logistic comp_case i.mated_binary, coef

gen logOR_X=e(b)[1,2]

logistic comp_case bwt_kg i.mated_binary, coef

gen logOR_W=e(b)[1,1]

keep if _n==1

/* Substituting in all terms gives: */

gen cov_XR=0.6*logOR_X*var_X

gen cov_WR=0.6*logOR_W*var_W

gen maxbias=(cov_XR*cov_WR*cov_YW)/((var_X - cov_XR^2)*var_W - (var_X*cov_WR^2))

di maxbias
